# Predicting Confrontation Naming in the Logopenic Variant of Primary Progressive Aphasia

**DOI:** 10.1101/2022.06.25.22276804

**Authors:** Fatima Jebahi, Katlyn Victoria Nickels, Aneta Kielar

## Abstract

Naming difficulties are prominent and pervasive in the logopenic variant of primary progressive aphasia (lvPPA) and are related to its underlying deficits in phonological processing. Importantly, some words appear to be more vulnerable to deterioration than others. We hypothesize that these differences can be explained, in part, by words’ unique psycholinguistic properties. Our study investigated the role of psycholinguistic properties of words, along with their underlying psycholinguistic factors, on confrontation naming performance in individuals with lvPPA. Naming accuracy data were collected from 10 individuals with lvPPA using the Boston Naming Test (BNT). For each test item, values were extracted for frequency, contextual diversity, age of acquisition (AoA), word length, phonological neighborhood density (PND), concreteness, semantic neighborhood density (SND), familiarity, arousal, and valence. We examined the effects of these psycholinguistic properties on naming accuracy using logistic regression analyses at the individual level and multiple linear regression analysis at the group level. Age of acquisition emerged as the strongest psycholinguistic predictor of naming accuracy in lvPPA at both the individual and group levels. Given that AoA and frequency are highly correlated, mediation analyses were performed to identify the relationships between AoA, frequency, and naming accuracy. The influence of AoA on naming accuracy was only partially mediated by frequency. Principal component analysis was performed to extract fundamental factors of the psycholinguistic properties. Four principal psycholinguistic factors were extracted. These were interpreted as “lexical-semantic usage”, “phonological simplicity”, “semantic disembodiment”, and “semantic pleasantness”. These factor scores were entered into multiple linear and logistic regression analyses to investigate their relative contribution to naming accuracy in lvPPA. Results indicated that “lexical-semantic usage”, “semantic disembodiment”, and “semantic pleasantness” predicted naming performance at the group level. Additionally, “lexical-semantic usage” and “semantic disembodiment” emerged as significant predictors at the individual level. The effects of the psycholinguistic properties and their factors and their theoretical implications are discussed in the context of phonological deficits in lvPPA and models of word naming.

**Highlights:**

– Age of acquisition (AoA) predicts naming in lvPPA
– The effect of AoA on naming corresponds with lvPPA’s phonological impairment
– The effect of AoA on naming is mostly direct and partially mediated by frequency
– PCA-extracted psycholinguistic factors independently predict naming in lvPPA
– Factors related to lexical semantics, semantics, and emotionality affect naming

## 1. Introduction

### 1.1. The Logopenic Variant of Primary Progressive Aphasia

Primary progressive aphasia (PPA) is a neurodegenerative syndrome characterized by progressive deterioration of language functions that result from atrophy of the predominantly left-lateralized language network. Classification of individuals into different PPA variants is based on cognitive-linguistic profiles and anatomical distributions of cortical atrophy (Gorno-Tempini et al., 2011; Mesulam et al., 2014; Montembeault, Brambati, Gorno-Tempini, & Migliaccio, 2018). Originally, PPA was broadly classified into fluent and non-fluent variants. According to the most recent classification criteria, these variants are referred to as semantic (svPPA) and agrammatic/non-fluent (nfvPPA) of PPA, respectively (Gorno-Tempini et al., 2011). A third variant, the logopenic variant of PPA (lvPPA), characterized by distinct neuropathology, marked patterns of cortical involvement, and a unique cognitive-linguistic profile (Henry & Gorno-Tempini, 2010; Henry et al., 2016), was formally described by Gorno-Tempini et al. (2011).

Neuropathological evidence suggests that lvPPA is most commonly associated with Alzheimer’s disease pathology, unlike svPPA and nfvPPA, which are more commonly associated with frontotemporal lobar degeneration (Giannini et al., 2017; Leyton et al., 2011; Mesulam et al., 2008, 2022; Rohrer et al., 2010). Further, cortical atrophy in lvPPA predominantly involves the left temporo-parietal brain regions, including posterior temporal cortex, inferior parietal lobule, temporoparietal junction, and in some cases can extend into the inferior frontal regions (GornoLJTempini et al., 2004; Henry & Gorno-Tempini, 2010; Leyton, Piguet, Savage, Burrell, & Hodges, 2012; Rogalski et al., 2014). Initially, cortical atrophy is strongly left-lateralized, and progression to the right hemisphere regions can be observed in later stages of the disease (Rohrer et al., 2013).

The disruption of the left temporo-parietal brain network in lvPPA has been linked to deficits in phonological processing and working memory (Henry & Gorno-Tempini, 2010; Henry et al., 2016). These deficits give rise to lexical retrieval difficulties, frequent word-finding pauses, and errors in speech sound selection that often result in phonological paraphasias (e.g., “frower” for “flower”; Henry & Gorno-Tempini, 2010). Indeed, a distinguishing characteristic of lvPPA is a marked deficit in naming, in the context of relatively spared semantic knowledge and syntactic and motor speech abilities. Although naming deficits are found in all three PPA variants, they are associated with phonological processing impairment in lvPPA (Henry et al., 2016; Leyton et al., 2015, 2012). Further, naming impairments in lvPPA may be accompanied by difficulties in repetition at the phrase and sentence levels, a manifestation of phonological loop dysfunction (Gorno-Tempini et al., 2011; Henry & Gorno-Tempini, 2010; Lukic et al., 2019; Mesulam et al., 2014). These difficulties are prominent and pervasive, giving rise to communication breakdowns that interfere with daily work and social activities, thereby significantly reducing individuals’ quality of life (Medina & Weintraub, 2007).

### 1.2. Stages of Naming

Previous research in clinical populations demonstrated that successful naming depends on phonological and semantic levels of processing (Henry, Beeson, Alexander, & Rapcsak, 2012; Lambon Ralph, Moriarty, & Sage, 2002; Lambon Ralph, Sage, & Roberts, 2000). At its core, naming is a complex cognitive task that involves several successive stages of processing. These stages include conceptual preparation, lexical selection, phonological code retrieval, phonological encoding, phonetic encoding, and articulation (Levelt, 2001). In the context of picture naming, the image of the object is converted into a percept that is linked to multimodal verbal and nonverbal representations. The first level where the percept is mapped onto a concept is known as the semantic level of linguistic processing and includes conceptual preparation and lexical selection stages of word naming. The second level of lexical retrieval is the phonological level, which consists of phonological word-form retrieval and phonological encoding. At this level, abstract phonological word forms are retrieved (i.e., phonological code retrieval, e.g., retrieving the one-syllable structure of /LJtriLJ/ for the concept of a “tree”) and sub-lexical phonological processes are mapped onto a word (i.e., phonological encoding, including phonological segmentation and syllabification). After that, these phonological representations undergo phonetic encoding, which finally lead to the generation of motor programs for articulation.

Some models of naming assume a unidirectional feed-forward serial process between these stages (e.g., Levelt, Roelofs, & Meyer, 1999), while other models assume an interactive bidirectional process (e.g., Dell, Chang, & Griffin, 1999). Regardless of directionality, there is general consensus that the above-mentioned stages are essential for successful word production and requisite both semantic and phonological levels of processing (Lambon Ralph et al., 2002). A breakdown in any of these levels of word processing can affect the successful execution of naming (Schwartz, Dell, Martin, Gahl, & Sobel, 2006). Therefore, while naming difficulty is a commonly observed manifestation in numerous disorders, it can be caused by breakdown at different stages of word retrieval. In the case of lvPPA, the breakdown is believed to be at the post-semantic phonological processing level (Henry et al., 2016; Leyton et al., 2015, 2012; Migliaccio et al., 2016). Although phonological processing is one of the main levels of word naming, it is not well understood how the breakdown at the phonological stages of word naming contributes to lexical retrieval difficulties in lvPPA.

### 1.3. Psycholinguistic Determinants of Naming Accuracy

Lexical retrieval difficulties in lvPPA are not uniform across all words. In fact, some words are named consistently more accurately than others. Examination of the psycholinguistic properties of words may provide insight as to why some words appear to be more susceptible to lexical retrieval difficulty than others. Current evidence suggests that the psycholinguistic properties of words influence the efficiency at which words are processed and named (Jurafsky, 2003; Wilson, Isenberg, & Hickok, 2009). These effects have been explored across a range of psycholinguistic properties, including frequency, contextual diversity, familiarity, age of acquisition (AoA), word length, phonological neighborhood density (PND), imageability and concreteness, semantic neighborhood density (SND), valence, and arousal (Adelman, Brown, & Quesada, 2006; Ellis, 2002; Juhasz, 2005; Lewellen, Goldinger, Pisoni, & Greene, 1993; Strain & Herdman, 1999; Vitevitch, Stamer, & Sereno, 2008). The strong effects of frequency on naming have been found consistently across different studies, with higher frequency words being more efficiently accessed than lower frequency words (Ellis, 2002; Kittredge, Dell, Verkuilen, & Schwartz, 2008). Further, evidence suggests that the contextual diversity of words, defined as the number of different contexts in which a word is experienced, is predictive of how accessible the word is, such that words that are more contextual diverse are more efficiently named than words with lower contextual diversity (Adelman et al., 2006; Pexman, Hargreaves, Siakaluk, Bodner, & Pope, 2008). Investigations of familiarity on word naming indicate that more familiar words are named faster than less familiar words (Lewellen et al., 1993). Furthermore, studies examining the effects of AoA provide robust evidence that earlier acquired words are retrieved more efficiently than those that are later acquired (Juhasz, 2005). Investigations of word length effects yield inconsistent conclusions (Pitt & Samuel, 2006; Vitevitch et al., 2008). Studies that examined the effects of PND, a measure of neighborhood size based on the number of words that differ from the target word by one phoneme, have shown inconsistent results (Vitevitch & Luce, 2016). In addition, words with high imageability and concreteness are associated with better performance on word naming tasks (Strain & Herdman, 1999). Concerning SND, a measure of neighborhood size based on number of words that are similar in meaning to the target word, evidence suggests that production is slower and less accurate for words with closer rather than more neighborhoods (Bormann, 2011; Fieder, Wartenburger, & Abdel Rahman, 2019; Mirman, 2011). More recently, influence of emotionality, particularly valence and arousal, on word processing have gained increasing attention (Crossfield & Damian, 2021; Kuperman, Estes, Brysbaert, & Warriner, 2014; Recio, Conrad, Hansen, & Jacobs, 2014). Although studies examining the effects of valence and arousal on word production have been scarce, evidence suggests that both valence and arousal can exert independent effects on word processing (Recio et al., 2014). Interestingly, the effects of valence on word processing were found to be mostly consistent, such that words associated with positive emotions, corresponding to higher valence scores, are recognized faster than negative words, while the effects of arousal were inconsistent across studies and accounted for a small amount of variance, when found significant (Kuperman et al., 2014; Recio et al., 2014).

The effects of some of these psycholinguistic properties on word naming have also been investigated in clinical populations, including individuals with post-stroke aphasia and svPPA (Alyahya, Halai, Conroy, & Lambon Ralph, 2018, 2020; Bastiaanse, Wieling, & Wolthuis, 2016; Gordon, 2002; Lambon Ralph, Graham, Ellis, & Hodges, 1998; Nickels & Howard, 1995). These investigations have used multiple linear regression analyses to determine the independent contributions of psycholinguistic properties on naming. Interestingly, the results are not homogeneous across populations, with certain psycholinguistic properties affecting naming performance in svPPA but not in post-stroke aphasia, and vice versa (Alyahya et al., 2020; Gordon, 2002; Lambon Ralph et al., 1998; Nickels & Howard, 1995). For example, significant effects of frequency and familiarity on naming were found in svPPA (Lambon Ralph et al., 1998) but not in post-stroke aphasia (Nickels & Howard, 1995). This distinction is possibly due to the differences in the nature and degree of involvement of phonological and semantic processing systems in naming across these clinical populations (Lambon Ralph et al., 2002; Levelt, 2001). Further, even when comparing studies that investigate the impact of psycholinguistic properties on naming within one clinical population, results are also often inconsistent (Bastiaanse et al., 2016; Lambon Ralph et al., 1998; Nickels & Howard, 1995).

Each of these psycholinguistic properties has been associated with different stages of word naming (Adelman et al., 2006; Alario et al., 2004; Ghasisin, Yadegari, Rahgozar, Nazari, & Rastegarianzade, 2015; Graves, Grabowski, Mehta, & Gordon, 2007; Hinojosa, Méndez-Bértolo, Carretié, & Pozo, 2010; Peramunage, Blumstein, Myers, Goldrick, & Baese-Berk, 2011; Wilson et al., 2009). Specifically, imageability and concreteness, arousal, valence have been related to the pre-linguistic semantic conceptual preparation stage (Alario et al., 2004; Hinojosa et al., 2010; Perret & Bonin, 2019). Familiarity and SND have been related to both conceptual preparation and lexical selection stages (Bormann, 2011; Ghasisin et al., 2015; Graves et al., 2007; Perret & Bonin, 2019; Wilson et al., 2009). Contextual diversity is thought to reflect lexical selection and phonological code retrieval stages (Adelman et al., 2006), whereas frequency and AoA have been linked to the phonological code retrieval stage of word naming.

Further, PND has been associated with stages of lexical selection, phonological code retrieval, phonological encoding, phonetic encoding, and articulation (Peramunage et al., 2011). Finally, word length has been most strongly associated with the stages from phonological encoding onwards (Alario et al., 2004; Graves et al., 2007; Perret & Bonin, 2019; Wilson et al., 2009).

Importantly, although psycholinguistic properties represent different word characteristics, many are strongly intercorrelated and their effects can be interactive (Adelman et al., 2006; Nickels & Howard, 1995; Reilly & Kean, 2007). In fact, there is a long-standing debate concerning the locus of the psycholinguistic effects on naming. For example, some propose that due to the high intercorrelations between AoA and frequency, AoA effects could be, at least partly, driven by frequency while others propose the opposite (Brysbaert & Ghyselinck, 2006; Gerhand & Barry, 1998; Ghyselinck, Lewis, & Brysbaert, 2004; Lewis, 1999). As high frequency words tend to be acquired earlier in life, and vice versa, their effects on naming can be hard to disentangle. This inherent intercorrelation between psycholinguistic properties makes it difficult to investigate the independent effects of these properties on naming. There is evidence suggesting that some psycholinguistic properties, such as AoA, may contribute to the observed effects of frequency on naming (Brysbaert & Ellis, 2016; Heikkola, Kuzmina, & Jensen, 2021). However, the degree to which AoA and frequency mediate naming in lvPPA has not been systematically investigated.

### 1.4. The Present Study

To date, the studies that have investigated the effects of psycholinguistic properties on picture naming in PPA have been conducted in individuals with svPPA (Bird, Lambon Ralph, Patterson, & Hodges, 2000; Hoffman, Meteyard, & Patterson, 2014; Lambon Ralph et al., 1998). However, these effects may not be applicable to lvPPA, given the differences in the lexical retrieval stage at which naming breakdown occurs between the two variants (Henry & Gorno-Tempini, 2010; Migliaccio et al., 2016). To the best of our knowledge, only one study has examined the effects of psycholinguistic properties on speech in lvPPA. The study investigated the influence of AoA, frequency, and orthographic neighborhood density on lexical decision and word recognition in lvPPA (Vonk et al., 2019). Thus, the influence of psycholinguistic properties on naming in lvPPA is still largely unknown.

In the present study, we extracted the psycholinguistic properties of 60 words that correspond to the items of the Boston Naming Test (BNT; (Kaplan, Goodglass, & Weintraub, 1983) and examined the influence of these properties on naming accuracy in lvPPA. The main aims were (1) to simultaneously explore the relative influence of ten psycholinguistic properties on naming accuracy in lvPPA, (2) to examine the mediating contributions of related psycholinguistic properties (AoA and frequency) on naming accuracy in lvPPA, and (3) to identify the core psycholinguistic factors using principal component analysis (PCA) and examine their contributions to naming accuracy in lvPPA.

## 2. Methods

### 2.1. Participants

Participants with lvPPA were recruited through The University of Arizona (UA) Speech, Language, and Hearing Clinic, Aphasia Research Project, and Banner University Medical Center Tucson. Prior to enrollment, all participants signed written informed consent approved by UA Institutional Review Board (IRB). Individuals with lvPPA were diagnosed by a neurologist or a speech-language pathologist based on speech-language evaluation and neurological exam. All participants completed medical case history questionnaire, underwent comprehensive neuropsychological and language testing, and completed a structural magnetic resonance imaging (MRI) scan. This information was used to confirm their clinical diagnosis based on consensus guidelines established by Gorno-Tempini et al. (2011). The study’s inclusion criteria included: (i) presentation of lvPPA in the absence of any other neurological or psychiatric disorder, (ii) received score greater than 0 on the BNT (Kaplan, Goodglass, & Weintraub, 1983), (iii) native English language proficiency, and (iv) normal or corrected-to-normal hearing and vision. Participants with a history of neurological conditions other than lvPPA were not included in the study. In addition, participants were not included if they presented with contraindications to MRI (e.g., the presence of a pacemaker, implanted device or electrodes, non-removable metal in the body). Demographic data are summarized in Table 1. The study included 10 participants with lvPPA (7 females, 3 males) with mean age of 68.80 (*SE* = 1.82). Their years of education had a mean of 16.30 (*SE* = 1.27). Their symptom time post onset ranged from 0 to 4 years (*M* = 1.80, *SE* = 0.47).

**Table 1:**
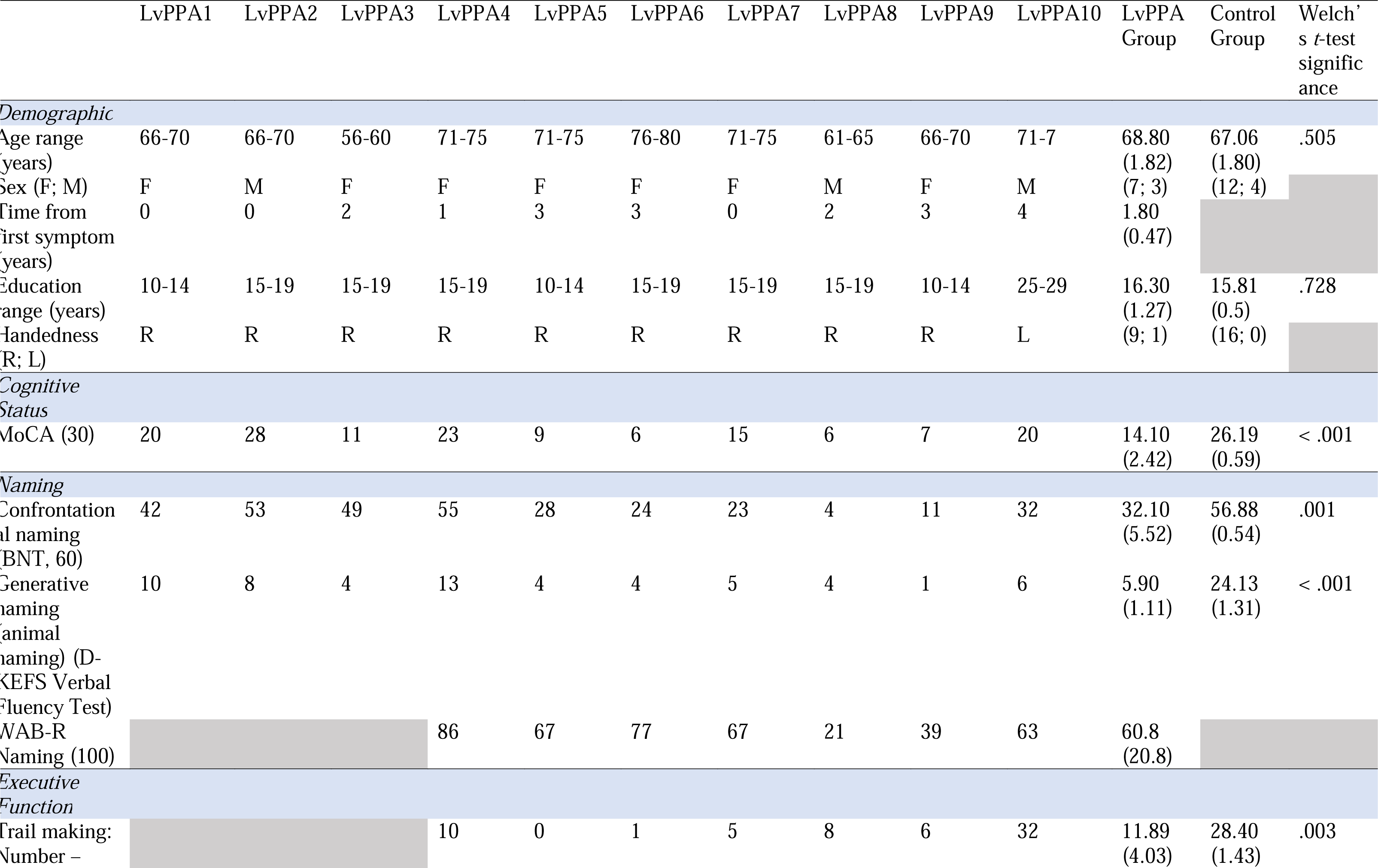
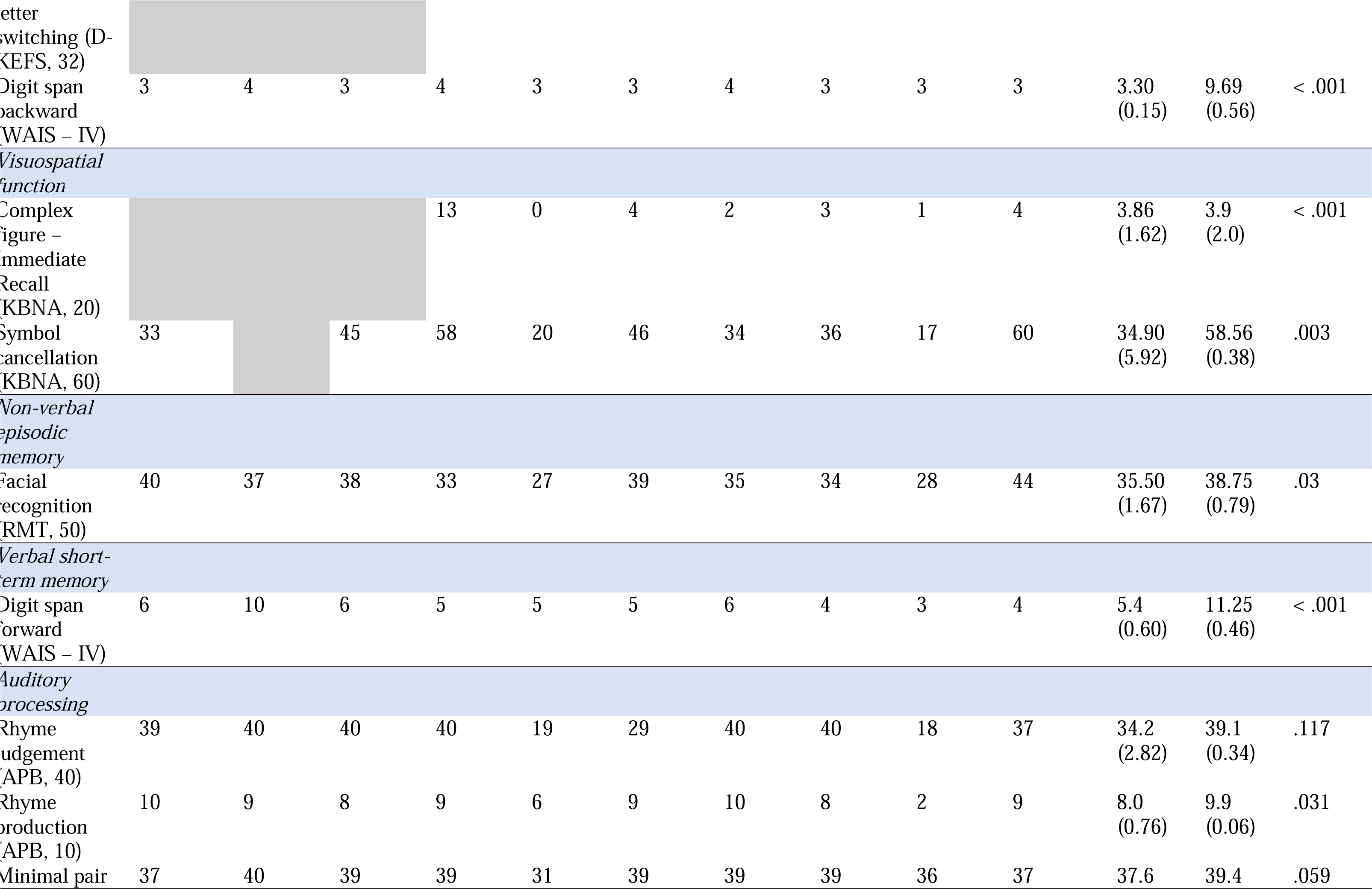
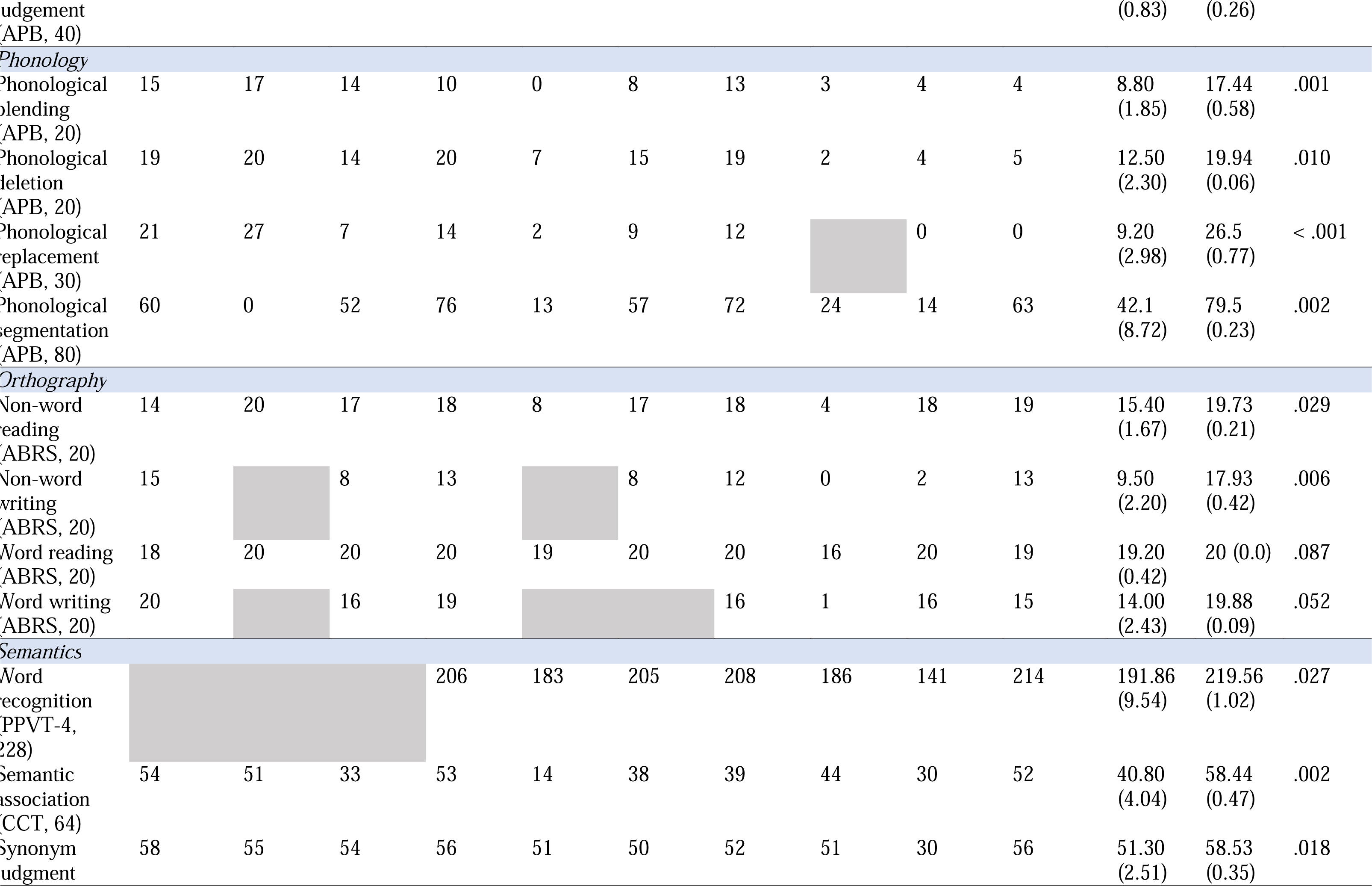
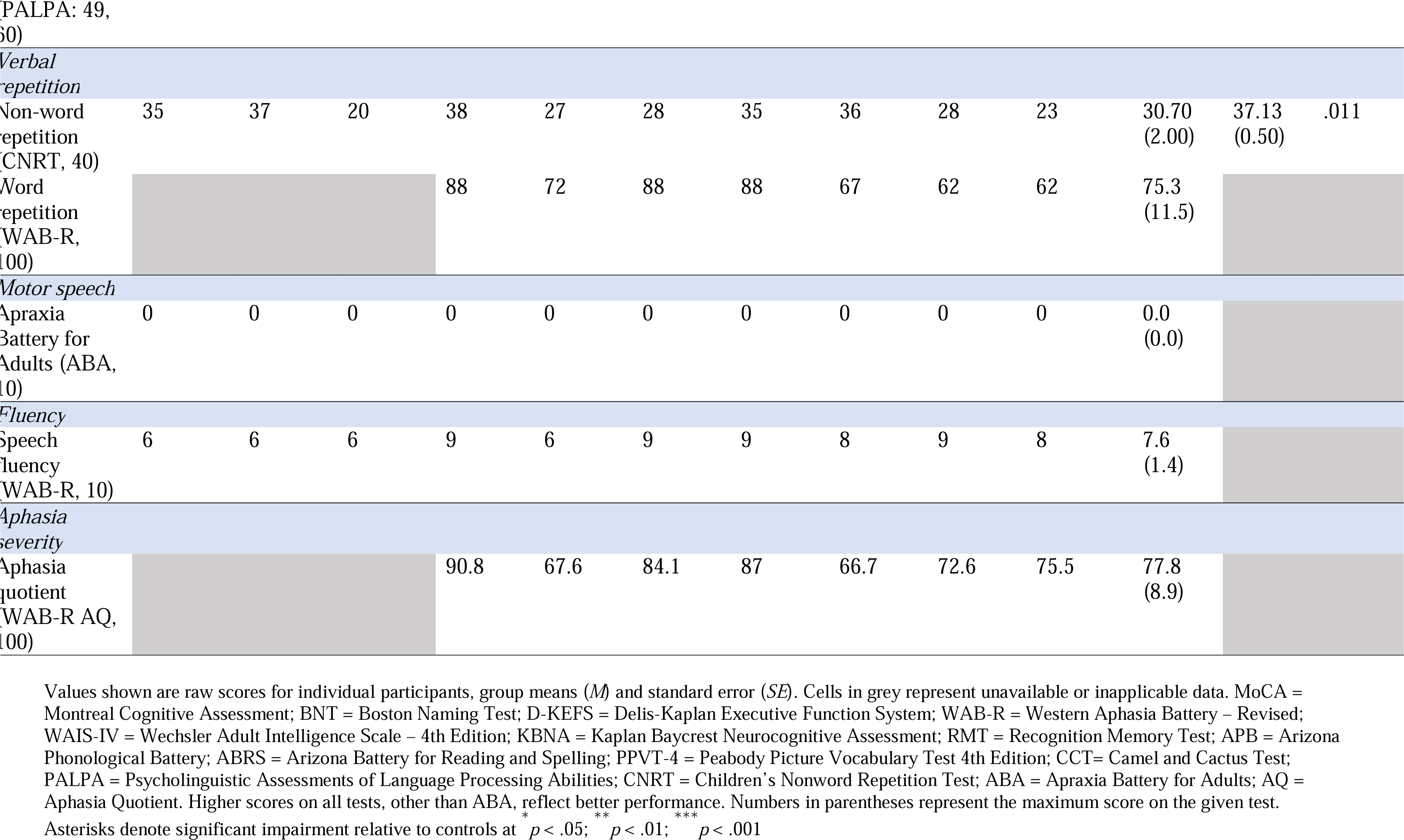
Demographic characteristics and neuropsychological data for individual lvPPA participants, lvPPA group, and control group

Participants with lvPPA were matched in age (*t*(22.34) = .678, *p* = .505) and education (*t*(11.84) = .356, *p* = .728) to a group of 16 neurologically unimpaired control participants. Control participants were recruited from the University of Arizona and Tucson community using IRB approved advertisements. All of the neurologically unimpaired participants (4 males, 12 females) were right-handed, native speakers of English, and had normal or corrected-to-normal hearing and vision. These participants had no history of neurological, psychiatric, speech, language, or learning disorders, and none were taking neuroleptic or mood-altering medications at the time of the study. They also underwent comprehensive neuropsychological and language testing, and completed a structural MRI scan, as those completed by the lvPPA participants. All control participants tested within normal limits on all behavioral tests (see Table 1).

### 2.2. Neuropsychological and Language Assessment

All participants completed a comprehensive cognitive and linguistic assessment battery to characterize their overall cognition, language performance, and sensorimotor skills. Scores for individual participants and group averages are presented in Table 1. Two lvPPA participants presented with sensorimotor deficits that prevented them from successfully completing all assessment tasks. Participant LvPPA8 had apraxic agraphia and an allographic impairment that precluded him from performing written language tasks. Participant LvPPA5 was also unable to complete written language tasks due to the presence of a tremor in her right hand.

Overall cognitive status was assessed using the Montreal Cognitive Assessment (MoCA; Nasreddine et al., 2005). Participants with lvPPA scored an average of 14.1 points, indicating moderate cognitive impairment across the group. Consistent with the logopenic subtype, participants demonstrated impairment on subtests of the MoCA that measured language skills (e.g., naming, sentence repetition), memory, and visuospatial function. Executive function was assessed using the digit span backwards (Wechsler Adult Intelligence Scale: WAIS-IV; Wechsler, 2008) and trail making test from Delis-Kaplan Executive Function System (D-KEFS; Delis, Kaplan, & Kramer, 2001). Visuospatial function was assessed using the complex figure copy and symbol cancellation tasks from the Kaplan–Baycrest Neurocognitive Assessment (KBNA; Leach, 2000) and nonverbal episodic memory was assessed using facial recognition from the Warrington Recognition Memory Test (RMT; Warrington, 1984). Motor-speech abilities were examined using the Apraxia Battery for Adults (ABA; Dabul, 2000), with higher scores indicating greater motor-speech impairment.

Quantifiable information on the relevant language skills of phonology, semantics, word retrieval, fluency, repetition, and comprehension is critical for differential diagnosis of PPA variants. Of particular importance is performance on phonological tasks, which has been shown to reliably predict lvPPA subtype (Henry et al., 2016; Leyton et al., 2014). To assess phonological skill, participants completed the Arizona Phonological Battery (ABP; Rapcsak & Beeson, 2004). Performance was markedly impaired on phoneme deletion, blending, replacement and segmentation subtests of APB (average accuracy = 48.6%). Participants also demonstrated a below-average digit span forward (Wechsler, 2008) score of 5.40 (*SE* = 0.60), indicative of the presence of phonological loop dysfunction. Written language was measured with the Arizona Battery for Reading and Spelling (ABRS; Beeson, Rising, Kim, & Rapcsak, 2010). The group demonstrated impaired nonword reading (77%) and nonword writing (48%), as compared to real word reading (96%) and writing (70%) tasks. This pattern is consistent with phonological alexia and agraphia profiles, which have been well-documented in individuals with lvPPA (Henry et al., 2016, 2012; Petroi et al., 2020).

Semantic skills were assessed using the Peabody Picture Vocabulary Test – 4th Edition (PPVT-4; Dunn & Dunn, 2007), the Camel and Cactus Test (CCT; Adlam, Patterson, Bozeat, & Hodges, 2010), and subtest 49 of the Psychologistic Assessment of Language Processing Abilities (PALPA; Kay, Lesser, & Coltheart, 1996). On average, participants demonstrated deficits on all semantic tasks, though performance was less impaired relative to phonological tasks.

Lexical retrieval difficulties, a hallmark characteristic of lvPPA, were also consistently captured by our assessment data. Confrontation naming was markedly impaired in all participants, with an average Boston Naming Test (BNT; (Kaplan et al., 1983) score of 32.1 (out of 60; *SE* = 5.5). Generative naming, measured by the D-KEFS verbal fluency task (Delis et al., 2001), was also below average. Finally, object naming was impaired on the Western Aphasia Battery – Revised (WAB-R; Kertesz, 2007) object naming subtest.

Another defining feature of lvPPA is impaired repetition, which is most pronounced at the phrase and sentence levels. Nonword repetition was measured using the Children’s Nonword Repetition Task (CNRT; Gathercole, Willis, Baddeley, & Emslie, 1994). Repetition of words, phrases, and sentences was measured with the WAB-R repetition subtest (Kertesz, 2007). Repetition performance was degraded on both the CNRT and WAB-R. Average overall aphasia severity, as measured by the WAB-R Aphasia Quotient (WAB-R AQ), was 77.6.

Overall, our cohort demonstrated cognitive-linguistic profiles consistent with the established diagnostic criteria for lvPPA, with some inherent heterogeneity across participants (Henry & Grasso, 2018; Louwersheimer et al., 2016). Participants with lvPPA demonstrated significantly impaired phonological skill (written and spoken), lexical retrieval, and repetition. Semantic impairment was also present in some participants; however, phonological deficits were more marked than semantic deficits. A subset of participants also demonstrated deficits in overall cognitive function including episodic memory, executive function, and visuospatial function. It is well-documented that individuals with lvPPA may experience degradation in these skills, especially in later stages of disease progression (Eikelboom et al., 2018; Kamath, Sutherland, & Chaney, 2020). In fact, episodic memory deficits in individuals with PPA are highly predictive of amyloid pathology, one of the hallmark pathological features of Alzheimer’s disease pathology, which is commonly observed in lvPPA (Ramanan et al., 2016). Thus, these cognitive-linguistic findings support the diagnosis of lvPPA.

### 2.3. Materials and Procedure

#### 2.3.1. Confrontation Naming

Picture naming was assessed using the BNT, a test consisting of 60 line-drawing object picture stimuli (Kaplan et al., 1983). The BNT is a widely used measure of confrontation naming in adults. Following standardized test administration, participants were shown one line-drawing picture at a time and asked to provide its name. Participants were given 20 seconds to spontaneously produce the name of the picture. Their responses were scored for accuracy according to the guidelines outlined by Nicholas, Brookshire, Maclennan, Schumacher, & Porrazzo (1989). Importantly, items on the BNT are ordered to gradually transition from more familiar to less familiar words (e.g., the item “bed” comes earlier than the item “abacus”).

#### 2.3.2. Extraction of Psycholinguistic Variables

Using the South Carolina Psycholinguistic Metabase (SCOPE; Gao, Shinkareva, & Desai, 2021), a collective repository of psycholinguistic databases, we extracted the values corresponding to seven psycholinguistic properties, including (1) Frequency, defined as the logarithm of base 10 of frequency norms based on the SUBTLEX_US_ corpus (Brysbaert & New, 2009); (2) Contextual Diversity, defined as the logarithm of base 10 of the number of passages in the SUBTLEX_US_ corpus containing the target word (Brysbaert & New, 2009); (3) AoA, defined as the subjective estimated age (in years) at which people acquired a given word (Kuperman, Stadthagen-Gonzalez, & Brysbaert, 2012), (4) Word length, was extracted from the English Lexicon Project (ELP; Balota et al., 2007) and represented by the number of phonemes in a given word; (5) PND, retrieved from ELP (Balota et al., 2007) and characterized by the mean logarithm hyperspace analogue to language (logHAL) of the phonological neighbors of a given word; (6) Concreteness, defined as the subjective degree to which the object can be experienced through the senses, with higher numbers indicating increased concreteness on a 5-point scale (Brysbaert, Warriner, & Kuperman, 2014); (7) SND, defined as the average distance between words in the semantic neighborhood and the target word (Shaoul & Westbury, 2010). Further, we extracted the values corresponding to three psycholinguistic properties from their respective individual databases, including (8) Familiarity, defined as the subjective experience pertaining to how familiar a word is, with higher numbers indicating greater familiarity on a 5-point scale (Himmanen, Gentles, & Sailor, 2003); (9) Valence, defined as the degree of emotional positivity and pleasantness associated with a given word, with scores ranging from 0 (extremely negative) to 1 (extremely positive) and higher scores indicating greater associated emotional pleasantness (Mohammad, 2018); and (10) Arousal, characterized by the degree of emotional intensity associated with a word, with scores varying between 0 (extremely calming) and 1 (extremely arousing) and higher scores indicating increased arousal (Mohammad, 2018).

### 2.4. Acquisition and Analysis of Neuroimaging Data

Participants underwent whole-brain T1-weighted structural MRIs using a 3T Siemens Magnetom Skyra MRI scanner located at the University of Arizona. The three-dimension magnetization-prepared rapid gradient echo (MP-RAGE) sequence was acquired with following parameters: 1 mm isotropic voxels, field-of-view = 256 mm, matrix = 256 x 256, 176 axial slices, repetition time (TR) = 2000 ms, time to echo (TE) = 2.33 ms, acquisition time (TA) = 293 s, scan time = 386 s. All images were acquired within one week of neuropsychological and language testing.

Voxel-based morphometry (VBM) implemented in Statistical Parametric Mapping – 12 (SPM12) software (Wellcome Department of Cognitive Neurology, London), was used to derive segmented, spatially normalized, bias corrected, and smoothed gray matter maps for all participants (Ashburner & Friston, 2005). Before processing, T1 images were evaluated for quality and were manually repositioned to set the anterior commissure as the origin to ensure consistent starting estimates for the unified segmentation routine. To increase the accuracy of inter-participant alignment, a nonlinear deformation toolbox was used (Ashburner, 2007). For each participant, flow fields were calculated during template creation that contained the nonlinear deformation information on the native image transformation to the template. These flow fields were applied to each participant’s image. The final template was registered to Montreal Neurological Institute (MNI) space using an affine transformation. This transformation was incorporated into the warping process, so that the individual spatially normalized scans could be brought into the common MNI space. During this final normalization step, the gray and white matter probability maps were scaled by their Jacobian determinants and smoothed using a 10 mm full width at half-maximum (FWHM) isotopic Gaussian kernel. To increase the accuracy of inter-participant alignment, nonlinear deformation parameters were calculated with the high dimensional diffeomorphic anatomical registration through exponentiated lie (DARTEL) algorithm and the predefined templates within the SPM12 DARTEL.

To identify cortical atrophy at the group level, the images of the lvPPA participants were compared with a group of 16 age- and education-matched controls using an independent sample t-test. Age, sex, education, and total intracranial volume were included as covariates. Specifically, we analyzed patterns of regional cortical atrophy in the brains of our lvPPA participants relative to the brains of the controls. The statistical maps were thresholded at voxel-wise threshold of *p* < .001 and corrected for multiple comparisons by controlling the family wise error (FWE) at the cluster level p < .05.

### 2.5. Analyses of Behavioral Data

The first aim was to examine influence of psycholinguistic properties on naming accuracy in lvPPA. To address this first aim, Pearson’s correlation coefficients were computed between naming accuracy and the extracted psycholinguistic values of BNT items, at the individual and group levels. The relative contributions of psycholinguistic properties to naming accuracy were investigated simultaneously with logistic regression analyses at the individual level and multiple linear regression analyses at the group level. In all regression analyses, naming accuracy on the BNT was entered as the dependent variable and the extracted psycholinguistic properties of words were entered as predictors.

To address our second aim, the mediating influence of related psycholinguistic properties on naming accuracy in lvPPA, we performed mediation analyses using model 4 of PROCESS macro (Hayes, 2017) in IBM SPSS Statistics (Version 28) analytics software with bootstrapping set to 5,000 samples and 95% confidence intervals. This analysis investigated whether the effect of AoA on naming accuracy in lvPPA can be explained by word frequency effect. Naming accuracy was entered as the outcome variable, AoA as the predictor, and frequency as the mediator.

To examine our third aim, we used PCA to identify core psycholinguistic dimensions and examine their contributions to naming accuracy in lvPPA. Principal component analysis is a multivariate data-driven decomposition approach that accounts for the maximum amount of shared variance in the data set and extracts principal components that can be used to derive fundamental structure of psycholinguistic properties. Values of all ten psycholinguistic properties for each BNT item were entered into the PCA and resulting factors with eigenvalues greater than 1 were extracted and varimax rotated. The PCA reduced the dimensionality of our large dataset of psycholinguistic properties, while preserving their unique characteristics. All input measures were standardized using functions in SPPS. The adequacy of the sample size for this PCA was determined using Kaiser-Meyer-Olkin measure of sampling adequacy and Bartlett’s test of sphericity. Finally, factor scores were extracted for each BNT item and entered into regression analyses as predictors of naming accuracy on BNT. These regression analyses were conducted at the individual and group levels.

## 3. Results

### 3.1. Voxel-based Morphometry

The results of the independent sample t-test comparing gray matter volumes in lvPPA versus age-matched control group are shown in Figure 1. Individuals with lvPPA showed areas of significant gray matter volume loss in the left hemisphere perisylvian language regions, including inferior parietal lobule (BA39/40), superior, middle, and inferior temporal gyri (BAs 22, 21, 20), and extended into the inferior (BA 44/45), and middle frontal gyri (BA8/9) and insula (BA13). Less extensive atrophy was found in the right hemisphere superior temporal gyrus (BA 22). These cortical atrophy patterns were consistent with those commonly seen in lvPPA, taking into account its neuroanatomical presentations, progression pattern, and individual heterogeneity (Henry & Gorno-Tempini, 2010; Leyton et al., 2015; Rohrer et al., 2013).

**Figure 1:**
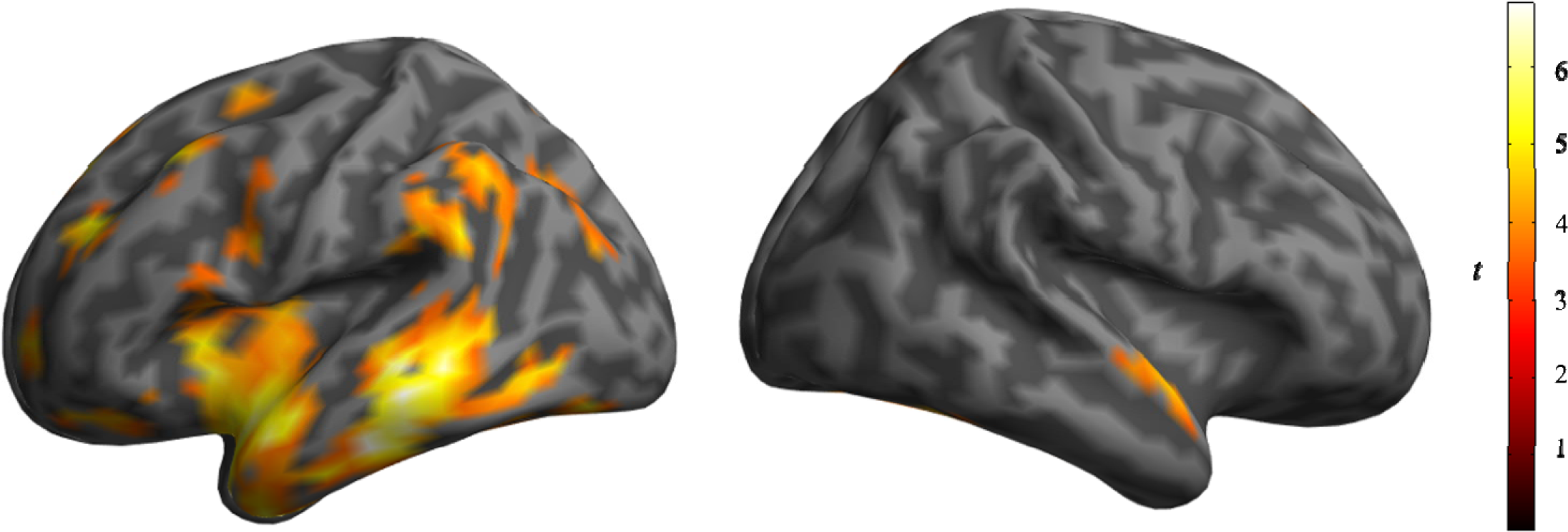
Statistical maps showing results of Voxel-based morphometry (VBM) analysis comparing gray matter volume in individuals with lvPPA (n=10) versus controls (n= 16) using independent samples t-test. The map shows regions of significant gray matter volume loss in lvPPA. The statistical maps were thresholded at voxel-wise threshold of p < .001 and corrected for multiple comparisons by controlling the family wise error (FWE) at the cluster level p < .05. Bright colors correspond to larger t-values and indicate greater atrophy in lvPPA relative to the matched control group

### 3.2. Behavioral Data

#### 3.2.1. Contributions of Psycholinguistic Properties to Naming Accuracy in LvPPA

The results of simple bivariate correlations between each of the psycholinguistic properties and naming accuracy for lvPPA participants at the individual and group levels are shown in Table 2. At the individual level, frequency and contextual diversity were significantly and positively correlated with naming accuracy in all but two lvPPA participants (LvPPA4 and LvPPA8). Age of acquisition was significantly and negatively correlated with naming accuracy for 7 lvPPA participants, indicating that earlier acquired words were named more accurately. Semantic neighborhood density was significantly and positively correlated with naming accuracy for 5 lvPPA participants, suggesting that words with more semantic neighbors were named more accurately. Additionally, familiarity was significantly and positively correlated with naming accuracy in 4 lvPPA participants. Concreteness was significantly and positively correlated with naming accuracy in 2 lvPPA participants (LvPPA1 and LvPPA3). In one lvPPA participant (lvPPA5), there was significant negative correlation between naming accuracy and word length and arousal, indicating that longer words and higher arousal words were named less accurately.

**Table 2:** Pearson’s correlations between psycholinguistic properties and naming accuracy for individual participants and LvPPA group

|  | LvPPA1 | LvPPA2 | LvPPA3 | LvPPA4 | LvPPA5 | LvPPA6 | LvPPA7 | LvPPA8 | LvPPA9 | LvPPA10 | LvPPA Group |
| --- | --- | --- | --- | --- | --- | --- | --- | --- | --- | --- | --- |
| Frequency | .384** | .392** | .387** | .153 | .456*** | .517*** | .265* | .251 | .543*** | .338** | .698*** |
| Contextual diversity | .381** | .366** | .387** | .149 | .465*** | .496*** | .293* | .223 | .528*** | .331** | .690*** |
| AoA | -.491*** | -.220 | -.533*** | -.176 | -.561*** | -.573*** | -.444*** | -.060 | -.554*** | -.332* | -.778*** |
| Word length | -.148 | -.195 | -.134 | .113 | -.413** | .179 | .073 | .076 | -.102 | -.128 | -.228 |
| PND | .024 | .083 | .108 | -.119 | .222 | .232 | .041 | -.089 | .110 | .001 | .150 |
| Concreteness | .279* | .145 | .267* | .121 | .263* | .080 | .115 | .106 | .055 | .212 | .311* |
| SND | .274* | .304* | .331** | .204 | .168 | .364** | .088 | .180 | .360** | .200 | .448*** |
| Familiarity | .312* | .219 | .523*** | .196 | .338** | .214 | .159 | .133 | .101 | .298* | .472*** |
| Arousal | -.101 | -.093 | -.120 | -.065 | -.331* | -.106 | -.109 | .047 | -.128 | -.151 | -.250 |
| Valence | .151 | -.093 | .235 | .195 | .165 | .213 | .204 | .119 | .183 | .123 | .289* |

At the group level, the analyses revealed significant positive correlations between naming accuracy and frequency, contextual diversity, concreteness, SND, familiarity, and valence. The relationship between naming accuracy and AoA was negative, indicating that earlier acquired words were named more accurately than words learned later in life. Correlations between naming performance, word length, PND and arousal were not significant.

To determine the relative contribution of each psycholinguistic property on naming accuracy, we used logistic and multiple linear regression analyses at the individual and group levels, respectively. Specifically, we performed a series of simultaneous forward-Wald logistic regression analyses at the individual level, and multiple linear regression analyses with all psycholinguistic properties at the group level. Results are presented in Table 3.

**Table 3:** Regression analyses of the effects of psycholinguistic properties on naming accuracy in individual participants and LvPPA group

|  | LvPPA1 | LvPPA2 | LvPPA3 | LvPPA4 | LvPPA5 | LvPPA6 | LvPPA7 | LvPPA8 | LvPPA9 | LvPPA10 | LvPPA Group |
| --- | --- | --- | --- | --- | --- | --- | --- | --- | --- | --- | --- |
| Frequency |  |  |  |  |  |  |  |  |  |  |  |
| $b$ (OR) / $B$ ($\beta$) | | | | | | | | | | | .190<br>(.605) |
| $p$ | n.s. | n.s. | n.s. | n.s. | n.s. | n.s. | n.s. | n.s. | n.s. | n.s. | .316 |
| Contextual diversity |  |  |  |  |  |  |  |  |  |  |  |
| $b$ (OR) / $B$ ($\beta$) | | | | | | | | | | | -.116<br>(-.329) |
| $p$ | n.s. | n.s. | n.s. | n.s. | n.s. | n.s. | n.s. | n.s. | n.s. | n.s. | .577 |
| AoA |  |  |  |  |  |  |  |  |  |  |  |
| $b$ (OR) / $B$ ($\beta$) | -.684<br>(.504) | | -1.169<br>(.311) | | -.888<br>(.411) | -.982<br>(.372) | -.554<br>(.574) | | -1.806<br>(.164) | -.342<br>(.710) | -.116<br>(-.502) |
| $p$ | .004** | n.s. | .010* | n.s. | < .001*** | < .001*** | .005** | n.s. | .002** | .046* | < .001*** |
| Word length |  |  |  |  |  |  |  |  |  |  |  |
| $b$ (OR) / $B$ ($\beta$) | | | | | -.417<br>(.659) | | | | | | .008<br>(.065) |
| $p$ | n.s. | n.s. | n.s. | n.s. | .035* | n.s. | n.s. | n.s. | n.s. | n.s. | .643 |
| PND |  |  |  |  |  |  |  |  |  |  |  |
| $b$ (OR) / $B$ ($\beta$) | | | | | | | | | | | -.003<br>(-.037) |
| $p$ | n.s. | n.s. | n.s. | n.s. | n.s. | n.s. | n.s. | n.s. | n.s. | n.s. | .790 |
| Concreteness |  |  |  |  |  |  |  |  |  |  |  |
| $b$ (OR) / $B$ ($\beta$) | | | | | | | | | | | .128<br>(.156) |
| $p$ | n.s. | n.s. | n.s. | n.s. | n.s. | n.s. | n.s. | n.s. | n.s. | n.s. | .113 |
| SND |  |  |  |  |  |  |  |  |  |  |  |
| $b$ (OR) / $B$ ($\beta$) | | | | | | | | | | | .009<br>(.005) |
| $p$ | n.s. | n.s. | n.s. | n.s. | n.s. | n.s. | n.s. | n.s. | n.s. | n.s. | .969 |
| Familiarity |  |  |  |  |  |  |  |  |  |  |  |
| $b$ (OR) / $B$ ($\beta$) | | | 5.688<br>(295.42) | | | | | | -3.886<br>(.021) | | .104<br>(.126) |
| *p* | n.s. | n.s. | .022<sup>*</sup> | n.s. | n.s. | n.s. | n.s. | n.s. | .038<sup>*</sup> | n.s. | .239 |
| Arousal |  |  |  |  |  |  |  |  |  |  |  |
| *b* (OR) / *B* ($\beta$) | | | | | | | | | | | .019<br>(.014) |
| *p* | n.s. | n.s. | n.s. | n.s. | n.s. | n.s. | n.s. | n.s. | n.s. | n.s. | .886 |
| Valence |  |  |  |  |  |  |  |  |  |  |  |
| *b* (OR) / *B* ($\beta$) | | | | | | | | | | | .090<br>(.057) |
| *p* | n.s. | n.s. | n.s. | n.s. | n.s. | n.s. | n.s. | n.s. | n.s. | n.s. | .569 |

At the individual level, the simultaneous forward-Wald logistic regression models were statistically significant (*p* ≤ .035) for all participants, except for LvPPA2, LvPPA4, and LvPPA8. Analyses revealed that AoA was a significant predictor of naming accuracy in seven participants: LvPPA1, LvPPA3, LvPPA5, LvPPA6, LvPPA7, LvPPA9, and LvPPA10. Familiarity was a significant predictor of naming accuracy in two participants: lvPPA3 and LvPPA9. Additionally, word length was a significant predictor for naming accuracy for LvPPA5.

At the group level, the regression model was significant (*R^2^* = .679, *F*(10,43) = 9.091, *p* < .001). The results indicated that, when controlling for all other psycholinguistic properties, AoA was the only psycholinguistic variable that significantly predicted naming accuracy (β = -.502, *p* < .001).

#### 3.2.2. Mediation Effects of Related Psycholinguistic Properties on Naming Accuracy in LvPPA

Results from the regression analyses indicated that AoA was the strongest predictor of naming accuracy in lvPPA at both the individual and group levels. However, frequency and AoA are strongly intercorrelated, such that earlier acquired words tend to be more frequent, and vice versa (Brysbaert & Ghyselinck, 2006; Morrison & Ellis, 1995). In order to investigate the extent to which the effect of AoA on naming accuracy may be driven by frequency, we performed mediation analysis. Using a series of multiple regressions, the mediation analysis assessed the indirect effect of AoA on the naming accuracy mediated by frequency, as well as the direct effect of AoA on naming accuracy, when the effect of frequency was controlled for. Results of mediation analysis are presented in Figure 2. These results suggest that the relationship between AoA and participants’ naming performance was partially (27.5% of the total effect) mediated by frequency (indirect effect = -.025, bootstrapping: *SE* = .009, 95% *CI* [-.042, -.008]). However, the direct effect of AoA on naming performance in our lvPPA sample accounted for most of the total effect (72.5% of the total effect) and remained statistically significant, even after frequency was included in the model (direct effect = -.066, *SE* = .012, 95% *CI* [-.090, -.042], *p* < .001). Consequently, the total effect of AoA on naming performance was mainly attributed to its direct effect and was partly mediated by frequency.

**Figure 2:**
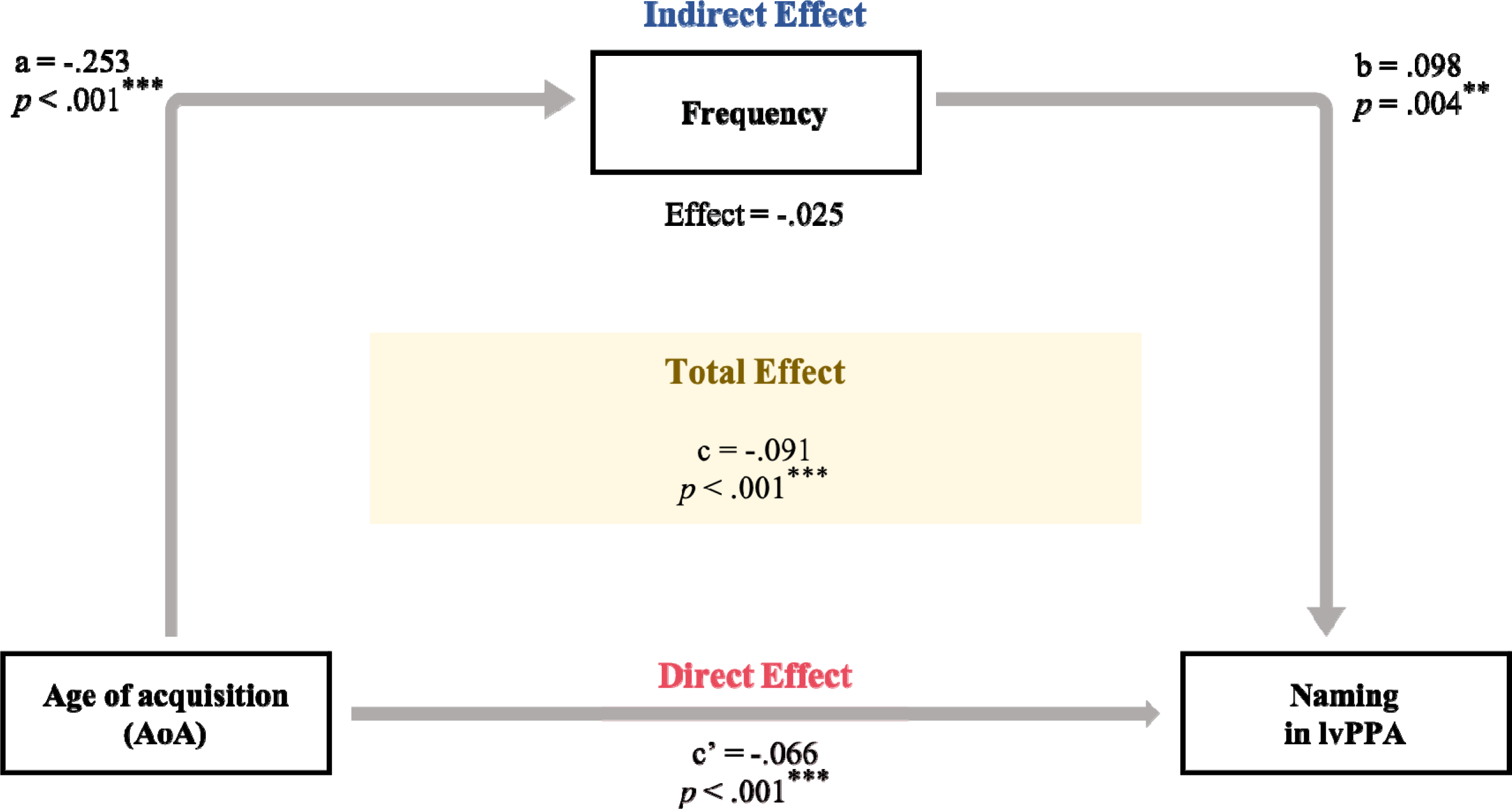
Mediation model with age of acquisition (AoA; predictor), frequency (mediator), and naming performance in lvPPA (outcome). Frequency partially mediated the relationship between AoA and naming performance in lvPPA; Asterisks denote significant effects on naming accuracy at **p <.01, ***p < .001; After frequency was included in the model, AoA was still a significant predictor of naming and accounted for 72.5% of the total effect of AoA on naming accuracy Frequency accounted for 27.5% of the total effect of AoA on naming

#### 3.2.3. Contributions of Psycholinguistic Factors to Naming Accuracy in LvPPA

Pearson correlation coefficients were computed to assess the linear relationship between each pair of psycholinguistic properties (frequency, contextual diversity, AoA, word length, PND, concreteness, SND, familiarity, arousal and valence). We presented these results in a correlation matrix, shown in Figure 3. The correlation matrix showed significant inherent intercorrelations between several our psycholinguistic properties.

**Figure 3:**
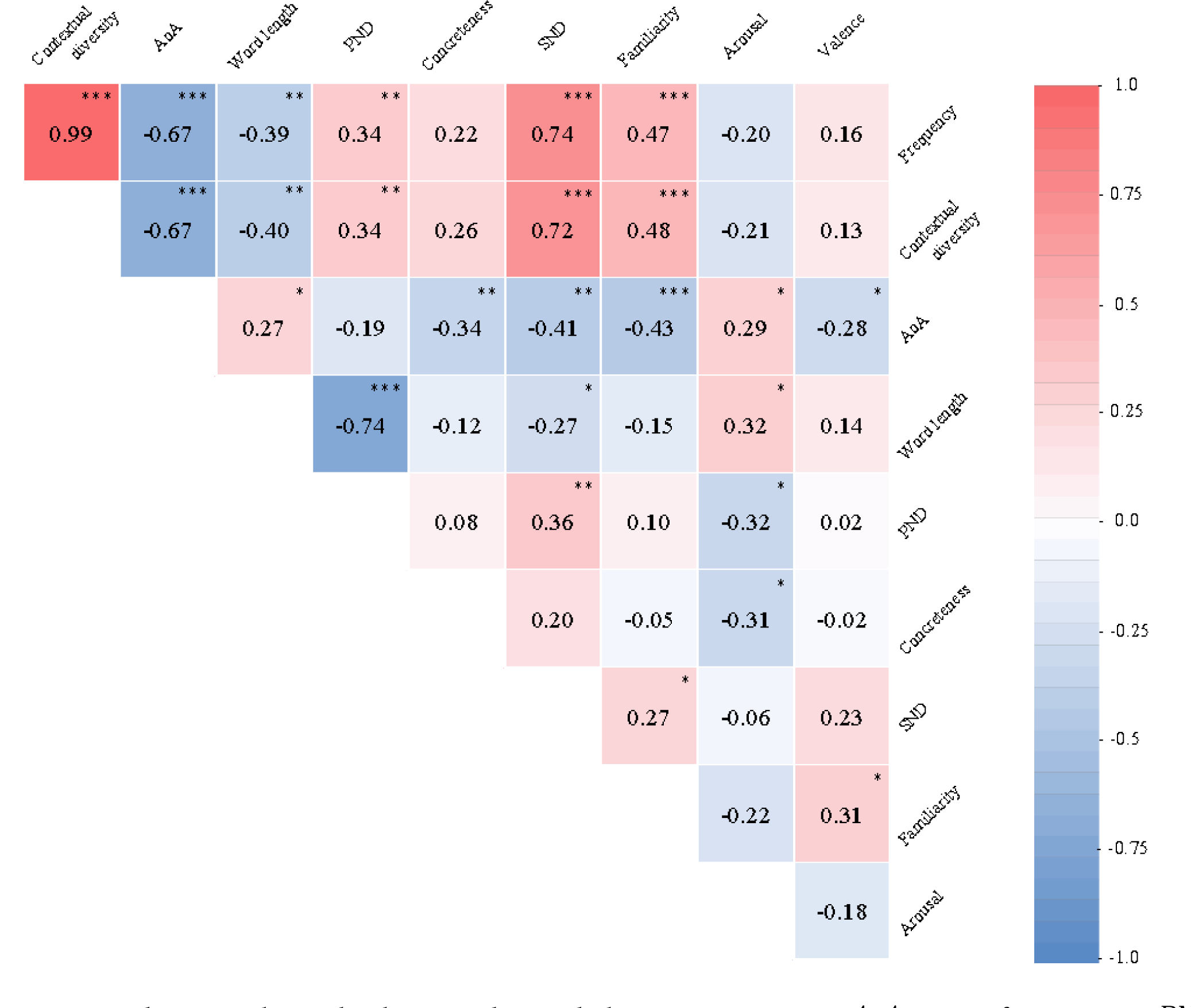
Correlation matrix showing relationship between the psycholinguistic properties; AoA = age of acquisition; PND = phonological neighborhood density; SND = semantic neighborhood density; Values in the correlation matrix represent Pearson’s correlation coefficient (r), Asterisks denote significant Pearson’s correlations at *p < .05, **p <.01, ***p < .001

In light of these findings, we explored the underlying structure of our extracted psycholinguistic properties using a varimax-rotated PCA. The PCA identified the underlying dimensions from our ten psycholinguistic properties, while accounting for their maximum amount of shared variance. Kaiser-Meyer-Olkin (KMO = .693) indicated that the sample size was acceptable and significant. Bartlett’s test of sphericity (χ^2^(45) = 341.71, *p* < .001) indicated that correlations between items were sufficiently large. Further, the varimax-rotated PCA produced four factors that summarized our ten extracted psycholinguistic properties, which accounted for 77.43% of the variance. The loadings of each psycholinguistic properties on each factor are shown in Table 4. The patterns of these loadings allowed for interpretation of the four psycholinguistic factors. The first factor was interpreted as “lexical-semantic usage” and received high positive loadings from frequency, familiarity, contextual diversity, SND, and a negative loading from AoA. Higher scores on this factor reflect words that have high frequency, familiarity, are found in multiple contexts, and are acquired earlier in life. Properties with highest loadings on the second factor were word length and PND, with a negative loading from word length and a positive loading from PND, indicating that shorter words have more phonological neighbors. Thus, this factor was interpreted as “phonological simplicity”. The third factor was named “semantic disembodiment” and received a high positive loading from arousal and a high negative loading from concreteness, indicating that more concrete words are less emotionally arousing. The fourth factor was termed as “semantic pleasantness” and it received high positive loadings from valence, reflecting words with positive emotional associations.

**Table 4:** Variance explained and loadings of psycholinguistic properties on factors extracted from varimax-rotated PCA

|  | Factor 1:<br>Lexical-semantic<br>usage | Factor 2:<br>Phonological<br>simplicity | Factor 3:<br>Semantic<br>disembodiment | Factor 4:<br>Semantic<br>pleasantness |
| --- | --- | --- | --- | --- |
| Variance explained (%) | 32.43 | 19.24 | 13.74 | 12.02 |
| Frequency | **.947** | .201 | -.040 | .014 |
| Contextual diversity | **.943** | .201 | -.078 | -.013 |
| AoA | **-.736** | .030 | .392 | -.171 |
| Word length | -.227 | **-.856** | .117 | .124 |
| PND | .138 | **.920** | .023 | .006 |
| Concreteness | .087 | -.020 | **-.862** | -.136 |
| SND | **.714** | .262 | .259 | .043 |
| Familiarity | **.560** | -.019 | -.074 | .392 |
| Arousal | .014 | -.430 | **.615** | -.417 |
| Valence | .119 | -.096 | .055 | **.899** |

To explore the effects of the psycholinguistic factors identified with PCA on naming accuracy in lvPPA, factors scores were extracted for each BNT item and were then entered as predictors into the regression models with naming accuracy as the dependent variable. Simultaneous forward-Wald logistic and multiple linear regression analyses were conducted at the individual and group levels, respectively. The results of these regression analyses are shown in Table 5.

**Table 5:** Results of regression analyses with PCA-extracted psycholinguistic factors as predictors of naming accuracy

|  | LvPPA1 | LvPPA2 | LvPPA3 | LvPPA4 | LvPPA5 | LvPPA6 | LvPPA7 | LvPPA8 | LvPPA9 | LvPPA10 | LvPPA Group |
| --- | --- | --- | --- | --- | --- | --- | --- | --- | --- | --- | --- |
| Lexical-semantic usage |  |  |  |  |  |  |  |  |  |  |  |
| *b* (OR) / *B* ($\beta$) | 1.917<br>(6.803) | | 3.943<br>(51.551) | | 1.416<br>(4.122) | 1.581<br>(4.859) | | | 1.509<br>(4.522) | .693<br>(2.000) | .153<br>(.708) |
| *p* | .003** | n.s. | .007** | n.s. | .003** | .001** | n.s. | n.s. | .002** | .041* | < .001*** |
| Phonological simplicity |  |  |  |  |  |  |  |  |  |  |  |
| *b* (OR) / *B* ($\beta$) | | | | | | | | | | | -.014<br>(-.065) |
| *p* | n.s. | n.s. | n.s. | n.s. | n.s. | n.s. | n.s. | n.s. | n.s. | n.s. | .449 |
| Semantic disembodiment |  |  |  |  |  |  |  |  |  |  |  |
| *b* (OR) / *B* ($\beta$) | -.835<br>(.434) | | -1.169<br>(.311) | | -1.498<br>(.224) | | | | | | -.069<br>(-.320) |
| *p* | .042* | n.s. | .036* | n.s. | .003** | n.s. | n.s. | n.s. | n.s. | n.s. | < .001*** |
| Semantic pleasantness |  |  |  |  |  |  |  |  |  |  |  |
| *b* (OR) / *B* ($\beta$) | | | | | | | | | | | .039<br>(.180) |
| *p* | n.s. | n.s. | n.s. | n.s. | n.s. | n.s. | n.s. | n.s. | n.s. | n.s. | .040* |

n.s. = not significant; *b* represents unstandardized regression weights of forward-Wald logistic regression and *B* represents unstandardized regression weights of multiple linear regression model; OR represents the odds ratio of logistic regression and $\beta$ represents the standardized regression weights of multiple linear regression; logistic regression analyses were used for individual participants and multiple linear regression analyses were used for the lvPPA group; Asterisks denote significant effects on naming accuracy at \* $p < .05$ , \*\* $p < .01$ , \*\*\* $p < .001$

At the individual level, the simultaneous forward-Wald logistic regression models were statistically significant for six participants. The analyses indicated that lexical-semantic usage was a strong and significant predictor for naming accuracy in LvPPA1, LvPPA3, LvPPA5, LvPPA6, LvPPA9 and LvPPA10. Additionally, semantic disembodiment emerged as a significant factor predicting naming accuracy in LvPPA1, LvPPA3, and LvPPA5. At the group level, the multiple linear regression model was statistically significant (*R^2^* = .640, *F*(4,49) = 21.79, *p* < .001). The analyses revealed that lexical-semantic usage, semantic disembodiment, and semantic pleasantness predicted naming accuracy in lvPPA. However, phonological simplicity did not emerge as a significant predictor.

## 4. Discussion

Naming difficulties in lvPPA are well-documented and represent one of the defining characteristics of the variant (Gorno-Tempini et al., 2011; Henry & Gorno-Tempini, 2010; Mesulam et al., 2014). However, these difficulties are not uniform across words, and some lexical items appear to be more vulnerable to deterioration. There is a large body of literature in unimpaired populations and individuals with post-stroke aphasia that document the influence of psycholinguistics properties on naming and lexical access (Alario et al., 2004; Alyahya et al., 2018; Hinojosa et al., 2010; Nickels & Howard, 1995; Wilson et al., 2009). Further, previous studies in svPPA found that successful word naming is influenced by psycholinguistic properties, such as frequency, AoA, and familiarity (Lambon Ralph et al., 1998). However, the relationship between words’ psycholinguistic properties and naming accuracy in lvPPA has not been explored. The main aim of the present study was to systematically examine the relative influence of psycholinguistic variables on naming accuracy in lvPPA.

### 4.1. Contributions of Psycholinguistic Properties to Naming Accuracy in LvPPA

Regression results indicated that AoA was the strongest psycholinguistic predictor of naming accuracy at both the individual and group levels. In particular, our results suggested that words acquired earlier in life are less susceptible to loss in lvPPA. The facilitative effect of earlier acquired words on naming accuracy has been documented across different clinical populations, including individuals with post-stroke aphasia, svPPA, and unclassified PPA (Brysbaert & Ellis, 2016; Lambon Ralph et al., 1998; Nickels & Howard, 1995; Ukita, Abe, & Yamada, 1999). Further, our results align with the arbitrary mapping hypothesis of AoA, which states that earlier acquired words have richer representations and are more strongly consolidated in the mental lexicon (Ellis & Lambon Ralph, 2000). This has been attributed to the high levels of neuroplasticity during childhood, at the time of their acquisition, making words learned earlier in life less susceptible to loss and deterioration (Brysbaert & Ellis, 2016; Chang, Monaghan, & Welbourne, 2019; Lambon Ralph & Ehsan, 2006).

Interestingly, the effect of AoA has been related to the post-semantic lexical retrieval level in several models of lexical access and naming (Alario et al., 2004; Barry, Morrison, & Ellis, 1997; Gerhand & Barry, 1998; Perret & Bonin, 2019). In particular, Barry et al. (1997) proposed that AoA influences the activation of phonological representations for word naming, after the semantic level and prior to the articulation level (see Figure 4). Our findings therefore provide further support to the notion that the breakdown of naming in lvPPA occurs at the post-semantic phonological level of processing (Henry et al., 2016; Leyton et al., 2015, 2012; Migliaccio et al., 2016). Figure 4 summarizes our findings in the context of stages of word naming and their potential relationship to the ten psycholinguistic properties and four psycholinguistic factors examined in our study based on findings from the literature.

**Figure 4:**
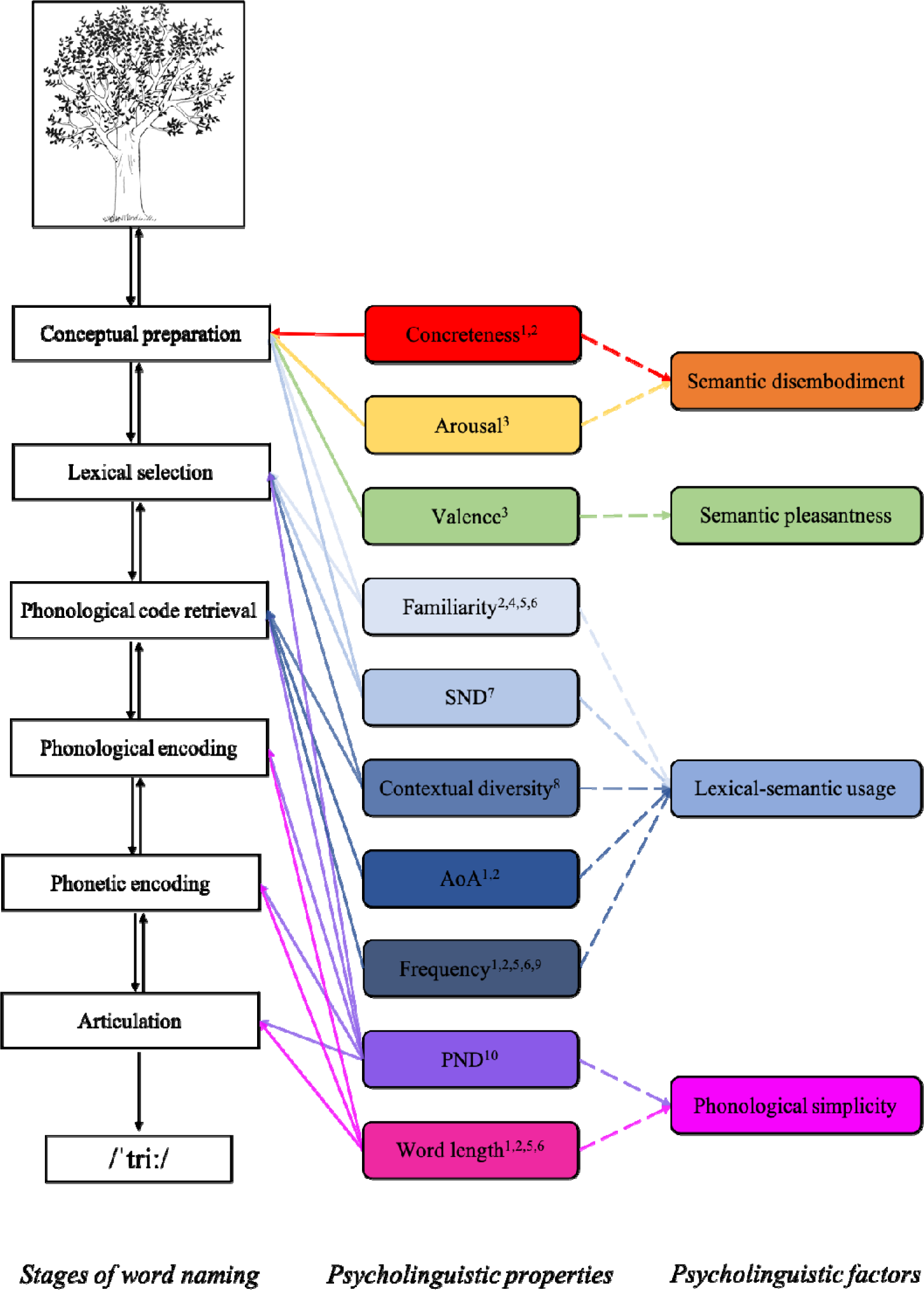
A graphical representation of the potential relationship between stages of word naming (Levelt, 2001), psycholinguistic properties and the underlying factors. The tree picture represents the second item on the Boston Naming Test (BNT; Kaplan et al., 1983). Colored solid line arrows represent mapping of psycholinguistic features on different stages of word naming based on ^1^Alario et al. (2004), ^2^Perret & Bonin (2019), ^3^Hinojosa et al. (2010), ^4^Ghasisin et al. (2015), ^5^Graves et al. (2007), ^6^Wilson et al. (2009), ^7^Bormann (2011), ^8^Adelman et al. (2006), ^9^Almeida et al. (2007), ^10^Peramunage et al. (2011). Dotted arrows represent loadings of psycholinguistic properties on PCA extracted factors. AoA = age of acquisition; PND = phonological neighborhood density; SND = semantic neighborhood density.

In addition to the effect of AoA, familiarity was a significant predictor of naming accuracy for two participants (LvPPA3 and LvPPA9), with more familiar words facilitating confrontation naming accuracy for these individuals. Familiarity has been related to semantic processing in models of naming, particularly to conceptual preparation leading to lexical selection (Ghasisin et al., 2015; Graves et al., 2007; Perret & Bonin, 2019; Wilson et al., 2009). Based on the results of cognitive and language assessment (see Table 1), LvPPA3 and LvPPA9 had pronounced semantic impairment, which is in line with their observed pattern of familiarity effect on naming accuracy. Moreover, adding to the effect of AoA, LvPPA5 demonstrated a significant effect of word length on naming accuracy, such that shorter words, consisting of fewer phonemes, were named more successfully than longer ones. Findings in the literature suggest that word length is related to phonological encoding, phonetic encoding, or articulation in models of naming (Alario et al., 2004; Graves et al., 2007; Perret & Bonin, 2019; Wilson et al., 2009). Notably, LvPPA5 exhibited pronounced receptive and expressive phonological impairment (see Table 1) and had basal ganglia involvement, all of which may explain her difficulty producing longer words (Silveri, 2021; Tettamanti et al., 2005). This suggests that more extensive phonological impairment in lvPPA5 may have affected several stages of word retrieval from phonological code retrieval to articulation. On the other hand, naming accuracy of participants LvPPA2, LvPPA4, and LvPPA8 was not significantly affected by any of our ten psycholinguistic properties. All three participants had limited variability in their naming performance, with LvPPA2 and LvPPA4 scoring very high (BNT score of 53 and 55 out of 60, respectively) and LvPPA8 scoring very low (BNT score of 4 out of 60) on the BNT. This limited variability may have reduced the impact of psycholinguistic properties on naming accuracy in these participants. Taken together, our results suggest that patterns of naming impairment in lvPPA can be explained by the psycholinguistic properties of words, and their interaction with underlying cognitive-linguistic abilities.

### 4.2. Mediation Effects of AoA and Frequency on Naming Accuracy in LvPPA

There is a long-standing debate in the literature concerning the relative contributions of AoA and word frequency on naming accuracy (Bonin, Chalard, Méot, & Fayol, 2002; Brysbaert & Ghyselinck, 2006; Ghyselinck et al., 2004; Morrison & Ellis, 1995). Specifically, it has been proposed that due to strong intercorrelations between these properties, such that words that are more frequent tend to be acquired earlier in life, and vice versa, the effects of AoA on naming could be, at least partly, influenced by frequency (for a review, see Ghyselinck et al., 2004). On the other hand, Bonin et al. (2002) suggested that when significant effects of frequency on naming are observed, they could be entirely attributed to AoA, such that influence of frequency becomes negligible when AoA is properly controlled for. Based on the results of our regression analyses, when all other psycholinguistic properties were controlled for, AoA emerged as the only psycholinguistic property affecting naming accuracy in lvPPA. To investigate the relative contribution of frequency on this effect, we performed a mediation analysis. The results suggested that the relationship between AoA and naming performance in lvPPA was partly mediated by frequency, but mostly attributed to the direct effects of AoA (see Figure 2). Therefore, the effects of AoA on naming in lvPPA appear to be only partly frequency-dependent and mostly frequency-independent, in line with previous findings (Brysbaert & Ghyselinck, 2006). Our findings are partially consistent with the arbitrary mapping hypothesis (Ellis & Lambon Ralph, 2000), indicating that words that are acquired earlier in life have richer and more stable representations.

### 4.3. Contributions of Psycholinguistic Factors to Naming Accuracy in LvPPA

The multiple linear and logistic regression analyses provided important information regarding the relative contributions of psycholinguistic properties to naming accuracy in lvPPA. However, the inherent intercorrelations between psycholinguistic properties (see in Figure 3) make it difficult to account for the unique variation of one psycholinguistic property while holding its highly correlated properties constant. To account for the shared variance among these properties, we used PCA, which is a multivariate data-driven decomposition method that simplifies the structure of high-dimensional data while retaining their patterns (Alyahya et al., 2018, 2020).

The PCA analysis yielded four psycholinguistic factors that underlie the structure of our ten psycholinguistic properties. The first factor, termed “lexical-semantic usage”, received high loadings from AoA, frequency, contextual diversity, SND, and familiarity, such that words that are more frequent, familiar, and contextually diverse had greater SNDs and were acquired earlier in life. The structure of this factor is similar to the “lexical usage” factor reported by Alyahya et al. (2020) and is consistent with studies that have reported the considerable inherent intercorrelations between its constituent psycholinguistic properties (Alyahya et al., 2020; Fergadiotis, Swiderski, & Hula, 2019; Lambon Ralph et al., 1998; Nickels & Howard, 1995). More importantly, its constituent properties have been associated with both semantic and lexical levels of processing in models of word naming (Adelman et al., 2006; Alario et al., 2004; Bormann, 2011; Ghasisin et al., 2015; Graves et al., 2007; Perret & Bonin, 2019; Wilson et al., 2009). The second factor obtained high loadings from word length (number of phonemes) and PND, such that shorter words had denser phonological neighbors. This factor was therefore termed “phonological simplicity”, and its structure is similar to that reported by Alyahya et al. (2020). In models of word naming, both word length and PND have been associated with stages from phonological code retrieval onward (Alario et al., 2004; Graves et al., 2007; Peramunage et al., 2011; Perret & Bonin, 2019; Wilson et al., 2009). The third factor was termed “semantic disembodiment” and received high loadings from concreteness and arousal, with the direction of loadings suggesting that less concrete words are more arousing. This relationship has been previously reported (Kousta, Vigliocco, Vinson, Andrews, & Del Campo, 2011) and follows on embodiment views of semantic representation (Kousta et al., 2011; Vigliocco et al., 2014). Interestingly, both concreteness and arousal have been associated with the conceptual preparation in models of word naming (Alario et al., 2004; Hinojosa et al., 2010; Perret & Bonin, 2019). Valence loaded heavily on the fourth factor that was named “semantic pleasantness”. In previous studies, valence has been associated with the semantic conceptual preparation stage in models of word naming, and conceptually, with emotional processing (Hinojosa et al., 2010). Figure 4 provides a graphical summary of the potential relationship between the stages of word naming according to Levelt (2001) and the psycholinguistic properties and their underlying factors examined in this study.

We examined the relative contribution of the PCA-extracted psycholinguistic factors on naming accuracy in our lvPPA sample. Findings from the multiple linear regression analyses revealed that “lexical-semantic usage”, “semantic disembodiment”, and “semantic pleasantness” factors were all independent predictors of naming performance in the lvPPA group. These findings suggest that words with greater lexical-semantic usage, lower semantic disembodiment, and greater semantic pleasantness tend to be named more accurately in lvPPA. Further, findings from the logistic regression analyses revealed that the lexical-semantic usage factor predicted naming accuracy in 6 individuals with lvPPA and the semantic disembodiment factor predicted naming accuracy in 3 individuals with lvPPA. Collectively, our findings align with previous studies that have provided evidence of the influence of lexical-semantic, semantic, and emotional processing on naming in clinical populations, albeit not lvPPA (Alyahya et al., 2020; Bormann, 2011; Jefferies & Lambon Ralph, 2006; Lampe, Hameau, Fieder, & Nickels, 2021; Newton, Thornley, & Bruce, 2020; Reilly et al., 2012). It is thus evident that the use of PCA factors, rather than individual psycholinguistic properties, as predictors reveals novel information that was previously masked, most likely due to the inherent intercorrelations that exist between psycholinguistic properties. The implementation of multivariate data-driven decomposition techniques in this study shed light on multidimensional effects of psycholinguistic properties on naming in lvPPA.

## 5. Conclusions

The findings of the current study suggest that psycholinguistic properties exert robust effects on naming accuracy in lvPPA. Age of acquisition emerged as the strongest psycholinguistic predictor of naming accuracy in lvPPA, at both the individual and group levels. The effect of AoA was mostly direct and only partially mediated by frequency. This pattern of psycholinguistic influence on naming in lvPPA is consistent with the underlying phonological impairment in lvPPA, given the well-established association of AoA with the phonological level of word naming. Clustering psycholinguistic properties into factors that underlie their structure and investigating them simultaneously revealed that lexical-semantic, semantic, and emotional processing independently affect word naming in lvPPA. Our findings offer important information regarding the organization of the lexical-semantic system and how it is affected by the phonological processing deficit found in lvPPA. The results of this study have clinical and empirical implications as they offer insights into the nature of naming deficits in lvPPA. Further, this study highlights the significance of word psycholinguistic properties and their underlying structures in naming, which can be informative in stimuli selection, manipulation, and interpretation in research and clinical practices. This information will be relevant for future studies to investigate the effects of psycholinguistic properties and factors on naming intervention outcomes in lvPPA.

## Data Availability

All data produced in the present study are available upon reasonable request to the authors

## Acknowledgments

This work was supported by the Arizona Alzheimer’s Consortium and by BIO5 Institute at the University of Arizona to author AK.

## CRediT authorship contribution statement

**Fatima Jebahi:** Conceptualization, Methodology, Investigation, Formal analysis, Visualization, Writing - Original Draft

**Katlyn Victoria Nickels:** Investigation, Visualization, Writing – Review & Editing

**Aneta Kielar:** Conceptualization, Formal analysis, Funding Acquisition, Resources, Supervision, Writing – Review & Editing

## References

1. Adelman, J. S., Brown, G. D. A., & Quesada, J. F. (2006). Contextual diversity, not word frequency, determines word-naming and lexical decision times. Psychological Science, 17(9), 814–823.

2. Adlam, A.-L. R., Patterson, K., Bozeat, S., & Hodges, J. R. (2010). The Cambridge Semantic Memory Test Battery: Detection of semantic deficits in semantic dementia and Alzheimer’s disease. Neurocase, 16(3), 193–207.

3. Alario, F., Ferrand, L., Laganaro, M., New, B., Frauenfelder, U. H., & Segui, J. (2004). Predictors of picture naming speed. Behavior Research Methods, Instruments, & Computers, 36(1), 140–155.

4. Alyahya, R. S. W., Halai, A. D., Conroy, P., & Lambon Ralph, M. A. (2018). Noun and verb processing in aphasia: Behavioural profiles and neural correlates. NeuroImage: Clinical, 18, 215–230.

5. Alyahya, R. S. W., Halai, A. D., Conroy, P., & Lambon Ralph, M. A. (2020). Mapping psycholinguistic features to the neuropsychological and lesion profiles in aphasia. Cortex, 124, 260–273.

6. Balota, D. A., Yap, M. J., Hutchison, K. A., Cortese, M. J., Kessler, B., Loftis, B., … Treiman, R. (2007). The English Lexicon Project. Behavior Research Methods, 39(3), 445–459.

7. Barry, C., Morrison, C. M., & Ellis, A. W. (1997). Naming the Snodgrass and Vanderwart pictures: Effects of age of acquisition, frequency, and name agreement. The Quarterly Journal of Experimental Psychology: Section A, 50(3), 560–585.

8. Bastiaanse, R., Wieling, M., & Wolthuis, N. (2016). The role of frequency in the retrieval of nouns and verbs in aphasia. Aphasiology, 30(11), 1221–1239.

9. Beeson, P. M., Rising, K., Kim, E. S., & Rapcsak, S. Z. (2010). A treatment sequence for phonological alexia/agraphia.

10. Bird, H., Lambon Ralph, M. A., Patterson, K., & Hodges, J. R. (2000). The rise and fall of frequency and imageability: Noun and verb production in semantic dementia. Brain and Language, 73(1), 17–49.

11. Bonin, P., Chalard, M., Méot, A., & Fayol, M. (2002). The determinants of spoken and written picture naming latencies. British Journal of Psychology, 93(1), 89–114. https://doi.org/10.1348/000712602162463

12. Bormann, T. (2011). The role of lexical-semantic neighborhood in object naming: implications for models of lexical access. Frontiers in Psychology, 2, 127.

13. Brysbaert, M., & Ellis, A. W. (2016). Aphasia and age of acquisition: are early-learned words more resilient? Aphasiology, 30(11), 1240–1263.

14. Brysbaert, M., & Ghyselinck, M. (2006). The effect of age of acquisition: Partly frequency related, partly frequency independent. Visual Cognition, 13(7–8), 992–1011. https://doi.org/10.1080/13506280544000165

15. Brysbaert, M., & New, B. (2009). Moving beyond Kučera and Francis: A critical evaluation of current word frequency norms and the introduction of a new and improved word frequency measure for American English. Behavior Research Methods, 41(4), 977–990.

16. Brysbaert, M., Warriner, A. B., & Kuperman, V. (2014). Concreteness ratings for 40 thousand generally known English word lemmas. Behavior Research Methods, 46(3), 904–911.

17. Chang, Y.-N., Monaghan, P., & Welbourne, S. (2019). A computational model of reading across development: Effects of literacy onset on language processing. Journal of Memory and Language, 108, 104025.

18. Crossfield, E., & Damian, M. F. (2021). The role of valence in word processing: Evidence from lexical decision and emotional Stroop tasks. Acta Psychologica, 218, 103359.

19. Dabul, B. (2000). Apraxia battery for adults: Examiner’s manual. Pro-ed.

20. Delis, D. C., Kaplan, E., & Kramer, J. H. (2001). Examiner’s Manual for the Delis-Kaplan Executive Function System. In Child Neuropsychology (Vol. 10). Retrieved from http://www.tandfonline.com/doi/abs/10.1080/09297040490911140

21. Dell, G. S., Chang, F., & Griffin, Z. M. (1999). Connectionist models of language production: Lexical access and grammatical encoding. Cognitive Science, 23(4), 517–542.

22. Dunn, L. M., & Dunn, D. M. (2007). PPVT-4: Peabody picture vocabulary test. Pearson Assessments.

23. Eikelboom, W. S., Janssen, N., Jiskoot, L. C., van den Berg, E., Roelofs, A., & Kessels, R. P. C. (2018). Episodic and working memory function in Primary Progressive Aphasia: A meta-analysis. Neuroscience & Biobehavioral Reviews, 92, 243–254.

24. Ellis, A. W., & Lambon Ralph, M. A. (2000). Age of Acquisition Effects in Adult Lexical Processing Reflect Loss of Plasticity in Maturing Systems: Insights from Connectionist Networks. Journal of Experimental Psychology: Learning Memory and Cognition, 26(5), 1103–1123. https://doi.org/10.1037/0278-7393.26.5.1103

25. Ellis, N. C. (2002). Frequency effects in language processing: A review with implications for theories of implicit and explicit language acquisition. Studies in Second Language Acquisition, 24(2), 143–188.

26. Fergadiotis, G., Swiderski, A., & Hula, W. D. (2019). Predicting confrontation naming item difficulty. Aphasiology, 33(6), 689–709.

27. Fieder, N., Wartenburger, I., & Abdel Rahman, R. (2019). A close call: Interference from semantic neighbourhood density and similarity in language production. Memory & Cognition, 47(1), 145–168.

28. Gao, C., Shinkareva, S. V., & Desai, R. H. (2021). SCOPE: The South Carolina Psycholinguistic Metabase. https://doi.org/10.31234/osf.io/mfkuq

29. Gathercole, S. E., Willis, C. S., Baddeley, A. D., & Emslie, H. (1994). The children’s test of nonword repetition: A test of phonological working memory. Memory, 2(2), 103–127.

30. Gerhand, S., & Barry, C. (1998). Word frequency effects in oral reading are not merely age-of-acquisition effects in disguise. *Journal of Experimental Psychology: Learning*, Memory, and Cognition, 24(2), 267.

31. Ghasisin, L., Yadegari, F., Rahgozar, M., Nazari, A., & Rastegarianzade, N. (2015). A new set of 272 pictures for psycholinguistic studies: Persian norms for name agreement, image agreement, conceptual familiarity, visual complexity, and age of acquisition. Behavior Research Methods, 47(4), 1148–1158.

32. Ghyselinck, M., Lewis, M. B., & Brysbaert, M. (2004). Age of acquisition and the cumulative-frequency hypothesis: A review of the literature and a new multi-task investigation. Acta Psychologica, 115(1), 43–67.

33. Giannini, L. A. A., Irwin, D. J., McMillan, C. T., Ash, S., Rascovsky, K., Wolk, D. A., … Grossman, M. (2017). Clinical marker for Alzheimer disease pathology in logopenic primary progressive aphasia. Neurology, 88(24), 2276–2284.

34. Gordon, J. K. (2002). Phonological neighborhood effects in aphasic speech errors: Spontaneous and structured contexts. Brain and Language, 82(2), 113–145.

35. Gorno-Tempini, M. L., Hillis, A. E., Weintraub, S., Kertesz, A., Mendez, M., Cappa, S. F., … Grossman, M. (2011). Classification of primary progressive aphasia and its variants. Neurology, 76(11), 1006–1014. https://doi.org/10.1212/WNL.0b013e31821103e6

36. GornoLJTempini, M. L., Dronkers, N. F., Rankin, K. P., Ogar, J. M., Phengrasamy, L., Rosen, H. J., … Miller, B. L. (2004). Cognition and anatomy in three variants of primary progressive aphasia. Annals of Neurology: Official Journal of the American Neurological Association and the Child Neurology Society, 55(3), 335–346.

37. Graves, W. W., Grabowski, T. J., Mehta, S., & Gordon, J. K. (2007). A neural signature of phonological access: distinguishing the effects of word frequency from familiarity and length in overt picture naming. Journal of Cognitive Neuroscience, 19(4), 617–631.

38. Hayes, A. F. (2017). Introduction to mediation, moderation, and conditional process analysis: A regression-based approach. Guilford publications.

39. Heikkola, L. M., Kuzmina, E., & Jensen, B. U. (2021). Predictors of object naming in aphasia: does cognitive control mediate the effects of psycholinguistic variables? Aphasiology, 1–18.

40. Henry, M. L., Beeson, P. M., Alexander, G. E., & Rapcsak, S. Z. (2012). Written language impairments in primary progressive aphasia: a reflection of damage to central semantic and phonological processes. Journal of Cognitive Neuroscience, 24(2), 261–275.

41. Henry, M. L., & Gorno-Tempini, M. L. (2010). The logopenic variant of primary progressive aphasia. Current Opinion in Neurology, 23(6), 633.

42. Henry, M. L., & Grasso, S. M. (2018). Assessment of individuals with primary progressive aphasia. Seminars in Speech and Language, 39(03), 231–241. Thieme Medical Publishers.

43. Henry, M. L., Wilson, S. M., Babiak, M. C., Mandelli, M. L., Beeson, P. M., Miller, Z. A., & Gorno-Tempini, M. L. (2016). Phonological processing in primary progressive aphasia. Journal of Cognitive Neuroscience, 28(2), 210–222.

44. Himmanen, S. A., Gentles, K., & Sailor, K. (2003). Rated familiarity, visual complexity, and image agreement and their relation to naming difficulty for items from the Boston Naming Test. Journal of Clinical and Experimental Neuropsychology, 25(8), 1178–1185.

45. Hinojosa, J. A., Méndez-Bértolo, C., Carretié, L., & Pozo, M. A. (2010). Emotion modulates language production during covert picture naming. Neuropsychologia, 48(6), 1725–1734.

46. Hoffman, P., Meteyard, L., & Patterson, K. (2014). Broadly speaking: Vocabulary in semantic dementia shifts towards general, semantically diverse words. Cortex, 55, 30–42.

47. Jefferies, E., & Lambon Ralph, M. A. (2006). Semantic impairment in stroke aphasia versus semantic dementia: a case-series comparison. Brain, 129(8), 2132–2147.

48. Juhasz, B. J. (2005). Age-of-acquisition effects in word and picture identification. Psychological Bulletin, 131(5), 684.

49. Jurafsky, D. (2003). Probabilistic modeling in psycholinguistics: Linguistic comprehension and production. Probabilistic Linguistics, 21.

50. Kamath, V., Sutherland, E. R., & Chaney, G.-A. (2020). A meta-analysis of neuropsychological functioning in the logopenic variant of primary progressive aphasia: Comparison with the semantic and non-fluent variants. Journal of the International Neuropsychological Society, 26(3), 322–330.

51. Kaplan, E., Goodglass, H., & Weintraub, S. (1983). Boston naming test.

52. Kay, J., Lesser, R., & Coltheart, M. (1996). Psycholinguistic assessments of language processing in aphasia (PALPA): An introduction. Aphasiology, 10(2), 159–180.

53. Kertesz, A. (2007). Western Aphasia Battery–Revised. San Antonio, TX: The Psychological Corporation.

54. Kittredge, A. K., Dell, G. S., Verkuilen, J., & Schwartz, M. F. (2008). Where is the effect of frequency in word production? Insights from aphasic picture-naming errors. Cognitive Neuropsychology, 25(4), 463–492.

55. Kousta, S.-T., Vigliocco, G., Vinson, D. P., Andrews, M., & Del Campo, E. (2011). The representation of abstract words: why emotion matters. Journal of Experimental Psychology: General, 140(1), 14.

56. Kuperman, V., Estes, Z., Brysbaert, M., & Warriner, A. B. (2014). Emotion and language: valence and arousal affect word recognition. Journal of Experimental Psychology: General, 143(3), 1065.

57. Kuperman, V., Stadthagen-Gonzalez, H., & Brysbaert, M. (2012). Age-of-acquisition ratings for 30,000 English words. Behavior Research Methods, 44(4), 978–990.

58. Lambon Ralph, M. A., & Ehsan, S. (2006). Age of acquisition effects depend on the mapping between representations and the frequency of occurrence: Empirical and computational evidence. Visual Cognition.

59. Lambon Ralph, M. A., Graham, K. S., Ellis, A. W., & Hodges, J. R. (1998). Naming in semantic dementia—what matters? Neuropsychologia, 36(8), 775–784.

60. Lambon Ralph, M. A., Moriarty, L., & Sage, K. (2002). Anomia is simply a reflection of semantic and phonological impairments: Evidence from a case-series study. Aphasiology, 16(1–2), 56–82.

61. Lambon Ralph, M. A., Sage, K., & Roberts, J. (2000). Classical anomia: A neuropsychological perspective on speech production. Neuropsychologia, 38(2), 186–202.

62. Lampe, L. F., Hameau, S., Fieder, N., & Nickels, L. (2021). Effects of semantic variables on word production in aphasia. Cortex, 141, 363–402.

63. Leach, L. (2000). The Kaplan–Baycrest neurocognitive assessment (KBNA): TEST manual. San Antonio, TX: Harcourt Assessment.

64. Levelt, W. J. M. (2001). Spoken word production: A theory of lexical access. Proceedings of the National Academy of Sciences, 98(23), 13464–13471.

65. Levelt, W. J. M., Roelofs, A., & Meyer, A. S. (1999). A theory of lexical access in speech production. Behavioral and Brain Sciences, 22(1), 1–38.

66. Lewellen, M. J., Goldinger, S. D., Pisoni, D. B., & Greene, B. G. (1993). Lexical familiarity and processing efficiency: individual differences in naming, lexical decision, and semantic categorization. Journal of Experimental Psychology: General, 122(3), 316.

67. Lewis, M. B. (1999). Age of acquisition in face categorisation: Is there an instance-based account? Cognition, 71(1), B23–B39.

68. Leyton, C. E., Hodges, J. R., McLean, C. A., Kril, J. J., Piguet, O., & Ballard, K. J. (2015). Is the logopenic-variant of primary progressive aphasia a unitary disorder? Cortex, 67, 122–133.

69. Leyton, C. E., Piguet, O., Savage, S., Burrell, J., & Hodges, J. R. (2012). The neural basis of logopenic progressive aphasia. Journal of Alzheimer’s Disease, 32(4), 1051–1059.

70. Leyton, C. E., Villemagne, V. L., Savage, S., Pike, K. E., Ballard, K. J., Piguet, O., … Hodges, J. R. (2011). Subtypes of progressive aphasia: application of the international consensus criteria and validation using β-amyloid imaging. Brain, 134(10), 3030–3043.

71. Louwersheimer, E., Keulen, M. A., Steenwijk, M. D., Wattjes, M. P., Jiskoot, L. C., Vrenken, H., … Scheltens, P. (2016). Heterogeneous language profiles in patients with primary progressive aphasia due to Alzheimer’s disease. Journal of Alzheimer’s Disease, 51(2), 581–590.

72. Lukic, S., Mandelli, M. L., Welch, A., Jordan, K., Shwe, W., Neuhaus, J., … Miller, B. L. (2019). Neurocognitive basis of repetition deficits in primary progressive aphasia. Brain and Language, 194, 35–45.

73. Medina, J., & Weintraub, S. (2007). Depression in primary progressive aphasia. Journal of Geriatric Psychiatry and Neurology, 20(3), 153–160.

74. Mesulam, M. M., Rogalski, E. J., Wieneke, C., Hurley, R. S., Geula, C., Bigio, E. H., … Weintraub, S. (2014). Primary progressive aphasia and the evolving neurology of the language network. Nature Reviews Neurology, 10(10), 554–569. https://doi.org/10.1038/nrneurol.2014.159

75. Mesulam, M, Coventry, C. A., Bigio, E. H., Sridhar, J., Gill, N., Fought, A. J., … Gefen, T. (2022). Neuropathological fingerprints of survival, atrophy and language in primary progressive aphasia. Brain.

76. Mesulam, Marsel, Wicklund, A., Johnson, N., Rogalski, E., Léger, G. C., Rademaker, A., … Bigio, E. H. (2008). Alzheimer and frontotemporal pathology in subsets of primary progressive aphasia. Annals of Neurology: Official Journal of the American Neurological Association and the Child Neurology Society, 63(6), 709–719.

77. Migliaccio, R., Boutet, C., Valabregue, R., Ferrieux, S., Nogues, M., Lehéricy, S., … Teichmann, M. (2016). The brain network of naming: a lesson from primary progressive aphasia. PloS One, 11(2), e0148707.

78. Mirman, D. (2011). Effects of near and distant semantic neighbors on word production. *Cognitive, Affective*, & Behavioral Neuroscience, 11(1), 32–43.

79. Mohammad, S. (2018). Obtaining reliable human ratings of valence, arousal, and dominance for 20,000 English words. Proceedings of the 56th Annual Meeting of the Association for Computational Linguistics (Volume 1: Long Papers), 174–184.

80. Montembeault, M., Brambati, S. M., Gorno-Tempini, M. L., & Migliaccio, R. (2018). Clinical, anatomical, and pathological features in the three variants of primary progressive aphasia: a review. Frontiers in Neurology, 692.

81. Morrison, C. M., & Ellis, A. W. (1995). Roles of word frequency and age of acquisition in word naming and lexical decision. *Journal of Experimental Psychology: Learning*, Memory, and Cognition, 21(1), 116.

82. Nasreddine, Z. S., Phillips, N. A., Bédirian, V., Charbonneau, S., Whitehead, V., Collin, I., … Chertkow, H. (2005). The Montreal Cognitive Assessment, MoCA: a brief screening tool for mild cognitive impairment. Journal of the American Geriatrics Society, 53(4), 695–699.

83. Newton, C., Thornley, H., & Bruce, C. (2020). The influence of emotional valence on word recognition in people with aphasia. Language, Cognition and Neuroscience, 35(8), 1064– 1072.

84. Nicholas, L. E., Brookshire, R. H., Maclennan, D. L., Schumacher, J. G., & Porrazzo, S. A. (1989). Revised administration and scoring procedures for the Boston Naming Test and norms for non-brain-damaged adults. Aphasiology, 3(6), 569–580.

85. Nickels, L., & Howard, D. (1995). Aphasic naming: What matters? Neuropsychologia, 33(10), 1281–1303.

86. Peramunage, D., Blumstein, S. E., Myers, E. B., Goldrick, M., & Baese-Berk, M. (2011). Phonological neighborhood effects in spoken word production: An fMRI study. Journal of Cognitive Neuroscience, 23(3), 593–603.

87. Perret, C., & Bonin, P. (2019). Which variables should be controlled for to investigate picture naming in adults? A Bayesian meta-analysis. Behavior Research Methods, 51(6), 2533– 2545.

88. Pexman, P. M., Hargreaves, I. S., Siakaluk, P. D., Bodner, G. E., & Pope, J. (2008). There are many ways to be rich: Effects of three measures of semantic richness on visual word recognition. Psychonomic Bulletin & Review, 15(1), 161–167.

89. Pitt, M. A., & Samuel, A. G. (2006). Word length and lexical activation: longer is better. Journal of Experimental Psychology: Human Perception and Performance, 32(5), 1120.

90. Ramanan, S., Flanagan, E., Leyton, C. E., Villemagne, V. L., Rowe, C. C., Hodges, J. R., & Hornberger, M. (2016). Non-verbal episodic memory deficits in primary progressive aphasias are highly predictive of underlying amyloid pathology. Journal of Alzheimer’s Disease, 51(2), 367–376.

91. Rapcsak, S. Z., & Beeson, P. M. (2004). The role of left posterior inferior temporal cortex in spelling. Neurology, 62(12), 2221–2229.

92. Recio, G., Conrad, M., Hansen, L. B., & Jacobs, A. M. (2014). On pleasure and thrill: The interplay between arousal and valence during visual word recognition. Brain and Language, 134, 34–43.

93. Reilly, J., & Kean, J. (2007). Formal distinctiveness of highLJand lowLJimageability nouns: Analyses and theoretical implications. Cognitive Science, 31(1), 157–168.

94. Reilly, J., Troche, J., Paris, A., Park, H., Kalinyak-Fliszar, M., Antonucci, S. M., & Martin, N. (2012). Lexicality effects in word and nonword recall of semantic dementia and progressive nonfluent aphasia. Aphasiology, 26(3–4), 404–427.

95. Rogalski, E., Cobia, D., Martersteck, A., Rademaker, A., Wieneke, C., Weintraub, S., & Mesulam, M.-M. (2014). Asymmetry of cortical decline in subtypes of primary progressive aphasia. Neurology, 83(13), 1184–1191.

96. Rohrer, J. D., Caso, F., Mahoney, C., Henry, M., Rosen, H. J., Rabinovici, G., … Fox, N. C. (2013). Patterns of longitudinal brain atrophy in the logopenic variant of primary progressive aphasia. Brain and Language, 127(2), 121–126.

97. Rohrer, J. D., Ridgway, G. R., Crutch, S. J., Hailstone, J., Goll, J. C., Clarkson, M. J., … Ourselin, S. (2010). Progressive logopenic/phonological aphasia: erosion of the language network. Neuroimage, 49(1), 984–993.

98. Schwartz, M. F., Dell, G. S., Martin, N., Gahl, S., & Sobel, P. (2006). A case-series test of the interactive two-step model of lexical access: Evidence from picture naming. Journal of Memory and Language, 54(2), 228–264.

99. Shaoul, C., & Westbury, C. (2010). Exploring lexical co-occurrence space using HiDEx. Behavior Research Methods, 42(2), 393–413.

100. Silveri, M. C. (2021). Contribution of the cerebellum and the basal ganglia to language production: Speech, word fluency, and sentence construction—evidence from pathology. The Cerebellum, 20(2), 282–294.

101. Strain, E., & Herdman, C. M. (1999). Imageability effects in word naming: an individual differences analysis. Canadian Journal of Experimental Psychology/Revue Canadienne de Psychologie Expérimentale, 53(4), 347.

102. Tettamanti, M., Moro, A., Messa, C., Moresco, R. M., Rizzo, G., Carpinelli, A., … Perani, D. (2005). Basal ganglia and language: phonology modulates dopaminergic release. Neuroreport, 16(4), 397–401.

103. Ukita, H., Abe, K., & Yamada, J. (1999). Late acquired words in childhood are lost earlier in primary progressive aphasia. Brain and Language, 70(2), 205–219.

104. Vigliocco, G., Kousta, S.-T., Della Rosa, P. A., Vinson, D. P., Tettamanti, M., Devlin, J. T., & Cappa, S. F. (2014). The neural representation of abstract words: the role of emotion. Cerebral Cortex, 24(7), 1767–1777.

105. Vitevitch, M. S., & Luce, P. A. (2016). Phonological neighborhood effects in spoken word perception and production. Annual Review of Linguistics, 2, 75–94.

106. Vitevitch, M. S., Stamer, M. K., & Sereno, J. A. (2008). Word length and lexical competition: Longer is the same as shorter. Language and Speech, 51(4), 361–383.

107. Vonk, J. M. J., Jonkers, R., Hubbard, H. I., Gorno-Tempini, M. L., Brickman, A. M., & Obler, L. K. (2019). Semantic and lexical features of words dissimilarly affected by non-fluent, logopenic, and semantic primary progressive aphasia. Journal of the International Neuropsychological Society, 25(10), 1011–1022.

108. Warrington, E. K. (1984). Recognition memory test. Western Psychological Services.

109. Wechsler, D. (2008). Wechsler adult intelligence scale–Fourth Edition (WAIS–IV). San Antonio, *TX*: *NCS Pearson*, 22(498), 816–827.

110. Wilson, S. M., Isenberg, A. L., & Hickok, G. (2009). Neural correlates of word production stages delineated by parametric modulation of psycholinguistic variables. Human Brain Mapping, 30(11), 3596–3608.

